# Genome-Wide Association Study of Creatinine Clearance Identifies New Loci for Kidney Function

**DOI:** 10.64898/2026.03.04.26347652

**Authors:** Anna D. Argoty-Pantoja, Peter J. van der Most, Zoha Kamali, Mahboube Ganji-Arjenaki, Amarens van der Vaart, Ahmad Vaez, Study Lifelines Cohort, Stephan J.L. Bakker, Harold Snieder, Martin H. de Borst

**Author notes:** These authors jointly supervised this work: Harold Snieder, PhD, Martin H. de Borst, MD, PhD. Lifelines Cohort Study contribution authors are listed in the supplemental material. **Corresponding Author:** Prof. Martin H. de Borst, MD, PhD. Department of Internal Medicine, Division of Nephrology. University Medical Center Groningen. P.O. Box 30.001, 9700 RB Groningen, The Netherlands.

## Abstract

**Introduction:** Genome-wide association studies (GWAS) for kidney function have mainly focused on creatinine-based glomerular filtration rate (eGFR_crea_), which is affected by variation in muscle mass. Moreover, the genetic basis of the sexual dimorphism of chronic kidney disease is underexplored.

**Methods:** We performed a GWA meta-analysis for creatinine clearance (CrCl), a muscle mass-independent kidney function phenotype, in 58,976 individuals of European descent from the Lifelines Cohort Study.

**Results:** We identified 16 independent loci with 21 genome-wide significant lead single nucleotide polymorphisms (SNPs) associated with CrCl, two of which had not been reported previously in kidney function GWASs: rs146465192, located near the *RP1-249F5.3* gene (effect allele frequency (EAF) = 0.01, *P* = 3.38 × 10^−9^) and rs117014836, located near the *AGPAT4* gene (EAF = 0.02, *P* = 5.42 × 10^−9^). Both SNPs were also associated with eGFR_crea_ in Lifelines (rs146465192: *P* = 1.34 × 10^−8^; rs117014836: *P* = 3.64 × 10^−7^), but not in previously published eGFR GWASs. *In silico* follow-up analyses revealed that rs146465192 was associated with plasma *IGF2R* (β = -0.519, *P* = 1.40 × 10^-6^), while rs117014836 was associated with blood expression levels of *AGPAT4* (eQTL *P* = 6.54 × 10^-6^). Furthermore, we identified two female-specific CrCl loci (t-statistic *P* < 0.004): rs8002366 (*GPC6*) and rs12908437 (*IGF1R*), associated with *GPC6* expression in kidney (eQTL *P* = 8.38 × 10^-10^) and *IGF1R* expression in blood (eQTL *P* = 2.62 × 10^-6^), respectively.

**Conclusion:** This first large-scale GWAS of CrCl revealed two new genetic variants among both sexes and two female-specific variants influencing kidney function.

**Lay summary:** Kidney function is a complex phenotype influenced by many different factors, including genetics. Earlier genetic studies often used the creatinine-based estimated glomerular filtration rate (eGFRcrea) as the measure of kidney function. However, eGFRcrea is influenced not just by kidney function but also by an individual’s muscle mass, which may distort the results. Therefore, in this study we used creatinine clearance (CrCl), a measure of kidney function independent of muscle mass, to look for genes in a European-ancestry population. We identified 16 genetic regions; two of which had not been found before. We also found two additional regions that were only related to CrCl in females. This shows the added value of investigating CrCl and suggests sex-based differences in how genetics affect kidney function.

## Introduction

Chronic kidney disease (CKD) is a significant global health issue, affecting >840 million people worldwide^1^. Risk factors include older age, male sex, lower socioeconomic status, hypertension, obesity, and diabetes. Genetic factors also contribute: heritability of estimated glomerular filtration rate (eGFR), the most commonly used method to estimate kidney function, is ∼45% in family studies^2^, and the most recent eGFR genome-wide association study (GWAS) estimated SNP-based heritability at 13.4%^3^. The gold standard for measuring kidney function involves the infusion and assessment of tracers such as inulin or iothalamate that are fully and exclusively cleared by the glomerulus^4^. However, in clinical practice, these complex, resource-intensive measurements are replaced by eGFR equations that most often use blood creatinine levels along with age and sex^5^. Despite its advantages, creatinine-based eGFR has limitations due to influences of non-GFR determinants such as muscle mass^6–8^, leading to under- or overestimation of kidney function. Creatinine clearance (CrCl) based on plasma creatinine and 24-hour urine creatinine excretion, overcomes this issue as it reflects kidney function independent of muscle mass.

Recent GWASs have made significant progress in elucidating the genetic variability of kidney function. A GWAS meta-analysis of creatinine-based eGFR identified 424 loci, collectively explaining 9.8% of the variance in eGFR^3^. Of note, kidney function displays sexual dimorphism^9–12^. While CKD is more prevalent in females, kidney failure is more common in males^10,13^. To date, little is known about the sex-specific genetic basis of kidney function.

Here, we perform GWA meta-analyses on CrCl as a muscle mass-independent kidney function phenotype in the Lifelines cohort, a large population-based study from the northern Netherlands, and conduct sex-specific GWAS analyses and bioinformatic follow-up analyses.

## Methods

### Study population

We used data from the Lifelines Cohort Study (UMCG Medical ethical committee under number 2007/152). Lifelines is a multi-disciplinary prospective population-based cohort study examining in a unique three-generation design the health and health-related behaviours of 167,729 persons living in the North of the Netherlands^14,15^. It employs a broad range of investigative procedures in assessing the biomedical, socio-demographic, behavioural, physical and psychological factors which contribute to the health and disease of the general population, with a special focus on multi-morbidity and complex genetics^14,15^. Participants ≥18 years at baseline were included, while individuals with missing phenotype data (n = 6,751) or inadequate 24-hour urine collections (i.e., over- or under-collection)^16^, defined as the upper or lower 2.5% of the difference between the estimated and measured 24-hour urine volume (n = 7,291), were excluded. Estimated 24-hour urine volume was derived from the creatinine clearance equation using the Cockcroft-Gault formula^17^. From 71,015 participants with genetic data, we removed overlapping genotyped samples and first-degree relatives (n = 12,010), and non-European samples (n = 29), resulting in a final sample of 58,976 participants (Supplementary Figure S1). We adhered to STROBE-STREGA guidelines^18^ (Supplementary Table S1).

### Phenotypes

The primary phenotype was CrCl, calculated as CrCl = (1000 *x* urine creatinine (mmol/l) * 24-hrs urine volume (ml)) / (serum creatinine (µmol/l) *x* 1440). CrCl was normalized to body surface area (BSA) (CrCl *x* 1.73 / sqrt (height *x* weight / 3600)) to account for differences in body composition and enable comparison with eGFR analyses^19^. The secondary outcome was estimated GFR (eGFR), calculated using the creatinine-based 2009 CKD Epidemiology Collaboration (CKD-EPI) equation^20^, to maintain consistency with prior GWAS^3^. Biochemistry methods were described previously^14,15^. Distributions of CrCl and eGFR were approximately normal (Supplementary Figures S2-S3), so no transformation were applied (Supplementary Methods).

### Genotyping, quality control and imputation

Genotyping was performed across three stages: CytoSNP, UGLI1, and UGLI2. CytoSNP used the Illumina CytoSNP-12v2 array^15^, UGLI1 the Infinium Global Screening Array® (GSA) MultiEthnic Disease Version^15^, and UGLI2 the FinnGen Thermo Fisher Axiom® custom array. Quality control and imputation were performed separately (Supplementary Table S2), as described previously^21,22^.

### Genome-wide association study (GWAS)

We performed GWASs for both CrCl and eGFR. Associations between each SNP and trait were tested assuming an additive model, using linear mixed models to account for relatedness within arrays^23,24^. Models were adjusted for age, sex (males/females), and the first twenty principal components (PCs) to control population stratification. Analyses were conducted with SAIGEgds (v1.12.5)^23^ for CytoSNP and UGLI1, and REGENIE (v3.4.1)^24^ for UGLI2. Both tools use linear mixed models for quantitative traits, account for sample relatedness, and have shown highly comparable association results^25–27^.

### Sex-specific genetic differences

We performed a sex-stratified GWAS adjusted for age and twenty PCs. For each genome-wide significant variant in either sex, we tested for sex differences using the t-statistics formula^28^:

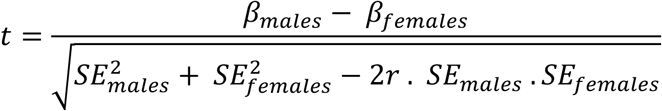

where r represents the Spearman correlation between β estimates across all sex-specific lead SNPs (n = 14).

A sex-difference was considered significant at the Bonferroni threshold (0.05/14 = 0.004). A SNP was considered a unique sex-specific SNP if it met the following three criteria: 1) genome-wide significant in one sex only, 2) not genome-wide significant in the sex-combined model, and 3) significant Bonferroni-corrected sex-difference through t statistic.

### Sensitivity analyses

We conducted two sensitivity analyses in sex-combined and sex-stratified models: (a) GWAS of CrCl unadjusted for BSA; and (b) GWAS of CrCl unadjusted for BSA with height included as a covariate.

### Quality control of GWAS results

Standardized quality control was performed using *GWASInspector* (v1.6.4)^29^, with 1000 Genomes Phase 3 (EUR) as reference^30^. We filtered out SNPs with minor allele frequency (MAF) <0.01, and imputation quality (INFO) <0.4. Genomic inflation was evaluated using QQ-plots and λ.

### Meta-analysis

Meta-analysis of CytoSNP, UGLI1, and UGLI2 was performed using fixed-effects inverse variance-weighting in METAL (v2020-05-05)^31^. This method weights the effect size estimates by their estimated standard errors^31^. The complete dataset comprised 7,791,514 markers for both CrCl and eGFR.

### Loci and significant SNPs

Genome-wide significant SNPs (*P* < 5 × 10^-8^) and loci were identified using the Functional Mapping and Annotation of GWAS platform (FUMA v1.5.2)^32^. Independent loci were pruned for linkage disequilibrium (LD) within a +/-250 kb window using the 1000 Genomes European population (EUR) LD reference panel. We selected independent significant lead SNPs with an r^2^ <0.1.

We compared locus coordinates from our GWAS results on CrCl and eGFR. Loci were considered overlapping if their genomic regions intersected.

### Comparison with eGFR_crea_ GWAS results

We examined associations between CrCl lead SNPs and the most recent creatinine-based eGFR GWAS in Europeans (n = 1,201,909) from the Chronic Kidney Disease Genetics consortium (CKDGen)^3^, comparing significance and consistency of effect direction, using publicly available summary statistics (https://ckdgen.imbi.uni-freiburg.de/). A similar lookup was performed using Lifelines eGFR_crea_ GWAS results from the present study.

### Genetic correlation and SNP-based heritability

We estimated genetic correlations (r_g_) between CrCl and eGFR_crea_ using LDSC v1.0 (https://github.com/bulik/ldsc/) with European LD scores from 1000 Genomes and HapMap3 SNPs^33,34^, and using GWAS summary statistics from CKDGen and Lifelines. SNP-based heritability was also estimated using LDSC with CKDGen data.

### Bioinformatics follow-up

#### In-silico lookup in GWAS Catalog

We explored functional characteristics of lead SNPs and surrounding regions (+/-500 kb) using *SNPannotator* (v0.2.6.0)^35^, which calculates LD (1Mb window, EUR reference) and selects SNPs with r^2^ ≥0.8 for downstream analyses.

#### Expression quantitative trait loci (eQTL) lookup

We searched blood and kidney eQTL databases. For blood, we used the cis- and trans-eQTL database from the eQTLGen consortium (n∼32,000)^36^. For kidney, we employed the kidney eQTL meta-analysis database (n = 686)^37^, and the TransplantLines kidney eQTL data (n∼200)^38^. We looked up novel SNPs and their highly linked proxies (r² >0.8), based on 1KG reference panel of European individuals.

#### Protein quantitative trait loci (pQTL) and Phenome-wide association studies (PheWAS) lookup

Novel lead SNPs identified in the CrCl GWAS were queried as look-ups in the Open Targets Genetics Platform (https://genetics.opentargets.org/), which enables the prioritization of candidate causal variants at trait-associated loci through publicly available data^39^. We searched for previously reported associations with protein expression via pQTLs and with specific traits through PheWAS, as reported in the original studies.

### Prioritization of kidney cell types for creatinine clearance

We prioritized kidney cell types associated with CrCl using scRNA-seq data from Stewart et al., which includes 33 annotated cell types from the cortex and medulla of 14 mature human kidneys^40^. Sex-combined GWAS summary statistics were processed using the munge_sumstats.py script from LDSC v1.0.0^33^, restricted to HapMap3 SNPs and excluding the MHC region (chr6:25-34Mb). Single-cell gene expression data were processed with CELLEX^41^ to derive expression specificity weights (ESw) and average expression specificity scores (ESµ). Cell type prioritization was evaluated using CELLECT^42^ with stratified LDSC to test cell-type enrichments for CrCl heritability. Cell-type-specific ESµ values were mapped to SNPs within a 100-kb window around gene transcription regions, LD scores were computed for GWAS SNPs, and stratified LDSC was used to estimate effect sizes and one-tailed *P* values for positive association of trait heritability and cell type ESµ values. Significance was assessed using Bonferroni-corrected and nominal thresholds. Additional methodological details are provided in the Supplementary Methods.

Additionally, we analysed *IGF2R, AGPAT4, IGF1R,* and *GPC6* expression using the renal scRNA-seq atlas^40^, with bar plots showing mean expression per cell type.

### Colocalization and fine-mapping analyses

To evaluate whether the same genetic variants underlie associations between creatinine clearance and gene expression or protein level, we performed colocalization analyses using the coloc package version 5.2.3^43^ and its SuSiE-based extension (coloc.susieR)^44^, and fine-mapping using FINEMAP v1.4.2^45^. These analyses were conducted for the two novel SNPs and the two female-specific SNPs identified in the GWAS (Supplementary Methods).

## Results

### Genome-wide association study meta-analysis

Phenotype characteristics are presented in Supplementary Table S3 (mean age 43.8 years; 60.4% female; mean CrCl 112 ml/min per 1.73m^2^, and mean eGFR 97 ml/min per 1.73m^2^). QQ-plots and λ are shown in Supplementary Figures S4-S5. For CrCl, across individual subcohorts, λ ranged from 0.95 to 1.15 for sex-combined and sex-stratified analyses. For the meta-analyses, λ values were 1.15, 1.09, and 1.06 for the sex-combined, female and male analyses, respectively.

The CrCl GWAS meta-analysis revealed 16 loci (Supplementary Table S4) with 21 lead SNPs significantly associated with CrCl (Figure 1, Table 1, Supplementary Table S5). Two novel SNPs, both on chromosome 6, had not been previously associated with kidney function (Table 1). The SNP rs146465192 (EAF = 0.01, *P* = 3.38 × 10^-9^) is located within a locus previously reported as genome-wide significant in CKDGen (rs3119304), with low LD between them (r^2^ = 0.050, EUR)^46,47^. The SNP rs117014836 (EAF = 0.02, *P* = 5.42 × 10^-9^) is located in a new locus (6:161421649-161632447). Hardy–Weinberg equilibrium values, and INFO scores for these SNPs across subcohorts are shown in Supplementary Table S6; locus zoom plots in Supplementary Figure S6. The two novel SNPs remained genome-wide significant after applying genomic inflation correction (Supplementary Table S7). Association statistics across individual Lifelines subcohorts for the two novel SNPs were consistent with the meta-analysis as indicated by limited heterogeneity between subcohorts (rs146465192: I² = 0% for total, female, and male; rs117014836: I² = 57.3% for total, 0% for female, and 55.15% for male) (Supplementary Figure S7, Supplementary Table S8).

**Figure 1.**
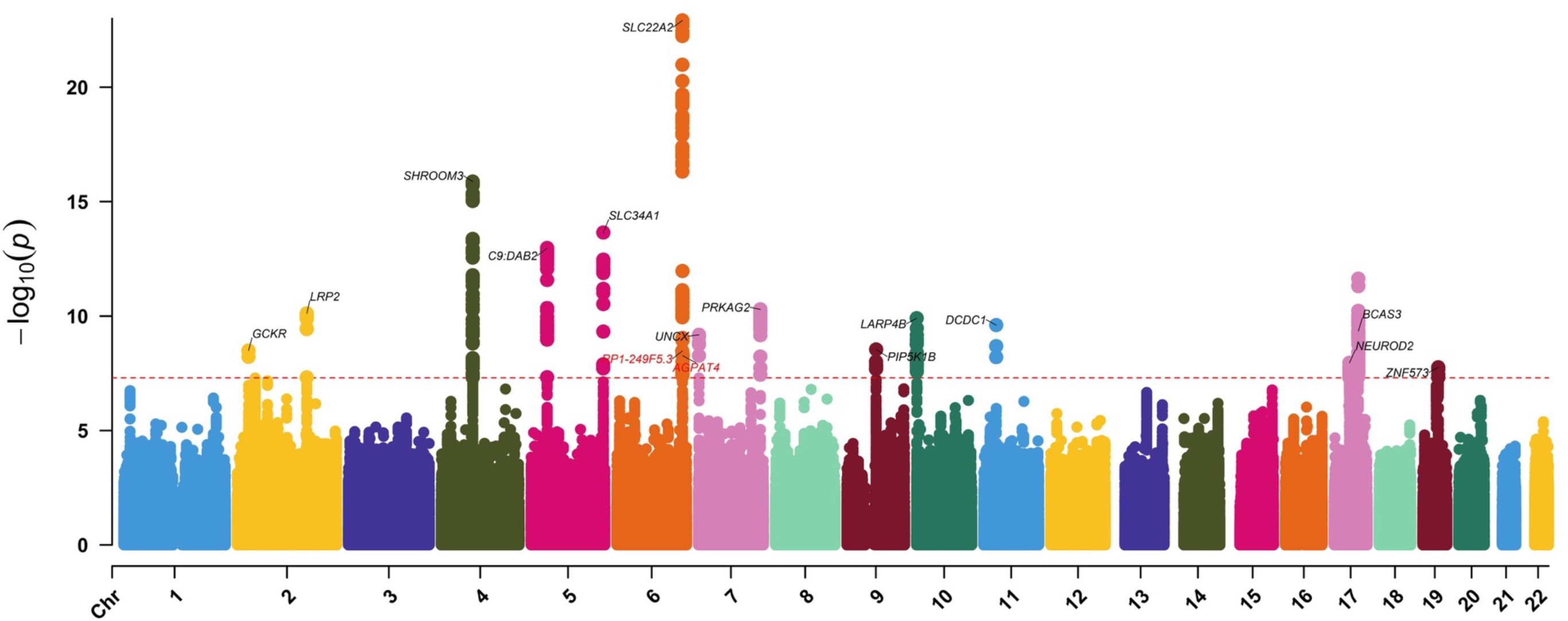
Manhattan plot for CrCl GWAS results. The Manhattan plot shows –log10 association P values for genetic effects on creatinine clearance by chromosomal base position (GRCh37). The red line indicates the threshold for genome-wide significance (p < 5 × 10⁻⁸). Models were adjusted for age, sex and 20 principal components (PCs). Gene names highlighted in red represent novel signals associated with kidney function. RP1-249F5.3 is the nearest gene to rs146465192 according to FUMA annotation; follow-up analyses identified this variant as a pQTL for *IGF2R* levels in blood.

**Table 1.** Overview of genome-wide significant lead SNPs for CrCl.

| Locus | SNP | Chr | Position | Type | Nearest gene | EA/<br>NEA | EAF | Effect | SE | P value | eGFR - CKDGen |  | eGFR - Lifelines |  |
| --- | --- | --- | --- | --- | --- | --- | --- | --- | --- | --- | --- | --- | --- | --- |
|  |  |  |  |  |  |  |  |  |  |  | Effect | P value | Effect | P value |
| 1 | rs1260326 | 2 | 27730940 | exonic | GCKR | T/C | 0.352 | 0.804 | 0.136 | $3.26 \times 10^{-9}$ | 0.005 | $1.84 \times 10^{-74}$ | 0.660 | $2.97 \times 10^{-19}$ |
| 2 | rs4140872 | 2 | 170026596 | intronic | LRP2 | T/C | 0.749 | -0.968 | 0.149 | $7.76 \times 10^{-11}$ | -0.003 | $9.73 \times 10^{-23}$ | -0.497 | $6.78 \times 10^{-10}$ |
| 3 | rs7674982 | 4 | 77412997 | intronic | SHROOM3 | A/G | 0.534 | -1.074 | 0.130 | $1.32 \times 10^{-16}$ | -0.007 | $6.93 \times 10^{-161}$ | -0.809 | $1.06 \times 10^{-30}$ |
| 4 | rs1362800 | 5 | 39378115 | intronic | C9:DAB2 | T/C | 0.418 | -0.970 | 0.131 | $1.08 \times 10^{-13}$ | -0.006 | $9.20 \times 10^{-101}$ | -0.654 | $2.10 \times 10^{-20}$ |
| 5 | rs78247455 | 5 | 176722005 | exonic | NSD1 | A/G | 0.037 | -1.941 | 0.341 | $1.29 \times 10^{-8}$ | -0.005 | $4.36 \times 10^{-09}$ | -0.804 | $1.36 \times 10^{-5}$ |
| 5 | rs3812036 | 5 | 176813404 | intronic | SLC34A1 | T/C | 0.249 | -1.145 | 0.150 | $2.24 \times 10^{-14}$ | -0.007 | $3.89 \times 10^{-109}$ | -0.789 | $2.56 \times 10^{-22}$ |
| **6** | **rs146465192** | **6** | **160297118** | **intergenic** | **RP1-249F5.3** | **A/C** | **0.014** | **-3.407** | **0.576** | **$3.38 \times 10^{-9}$** | **-0.004** | **0.0102** | **-1.775** | **$1.34 \times 10^{-8}$** |
| 7 | rs2279463 | 6 | 160668389 | intronic | SLC22A2 | A/G | 0.877 | 1.892 | 0.198 | $1.04 \times 10^{-21}$ | 0.008 | $2.46 \times 10^{-94}$ | 1.137 | $2.33 \times 10^{-26}$ |
| 7 | rs316035 | 6 | 160696544 | intronic | SLC22A2:RP1-276N6.2 | T/C | 0.159 | 1.768 | 0.176 | $1.21 \times 10^{-23}$ | 0.007 | $2.59 \times 10^{-67}$ | 1.074 | $2.38 \times 10^{-29}$ |
| **8** | **rs117014836** | **6** | **161516388** | **intronic** | **MAP3K4:AGPAT4** | **A/G** | **0.983** | **2.914** | **0.500** | **$5.42 \times 10^{-9}$** | **0.004** | **0.0023** | **1.375** | **$3.64 \times 10^{-7}$** |
| 9 | rs62435145 | 7 | 1286567 | intergenic | UNCX | T/G | 0.696 | -0.881 | 0.143 | $6.62 \times 10^{-10}$ | -0.006 | $1.83 \times 10^{-88}$ | -0.688 | $5.70 \times 10^{-19}$ |
| 10 | rs10480299 | 7 | 151405818 | intronic | PRKAG2 | T/C | 0.747 | 0.972 | 0.148 | $5.07 \times 10^{-11}$ | 0.008 | $1.73 \times 10^{-134}$ | 0.777 | $2.82 \times 10^{-22}$ |
| 11 | rs1411992 | 9 | 71170702 | intronic | RP11-274B18.4 | C/G | 0.81 | -0.943 | 0.164 | $9.85 \times 10^{-9}$ | -0.003 | $4.45 \times 10^{-20}$ | -0.548 | $7.37 \times 10^{-10}$ |
| 11 | rs4744712 | 9 | 71434707 | intronic | PIP5K1B | A/C | 0.401 | -0.785 | 0.132 | $2.90 \times 10^{-9}$ | -0.005 | $1.26 \times 10^{-67}$ | -0.594 | $1.04 \times 10^{-16}$ |
| 12 | rs35772020 | 10 | 863482 | intronic | LARP4B | A/G | 0.085 | -1.489 | 0.232 | $1.25 \times 10^{-10}$ | -0.008 | $9.53 \times 10^{-55}$ | -0.913 | $3.27 \times 10^{-13}$ |
| 13 | rs3925584 | 11 | 30760335 | intergenic | DCDC1 | T/C | 0.575 | -0.848 | 0.134 | $2.47 \times 10^{-10}$ | -0.005 | $9.31 \times 10^{-86}$ | -0.593 | $3.24 \times 10^{-16}$ |
| 14 | rs4795386 | 17 | 37746847 | intergenic | NEUROD2 | A/G | 0.289 | 0.828 | 0.145 | $1.11 \times 10^{-8}$ | 0.005 | $5.51 \times 10^{-61}$ | 0.665 | $2.55 \times 10^{-17}$ |
| 15 | rs11868441 | 17 | 59239221 | intronic | BCAS3 | A/G | 0.211 | 0.987 | 0.158 | $4.57 \times 10^{-10}$ | 0.006 | $1.55 \times 10^{-64}$ | 0.711 | $1.08 \times 10^{-16}$ |
| 15 | rs11657044 | 17 | 59450105 | ncRNA_intronic | BCAS3:RP11-332H18.5 | T/C | 0.152 | -1.290 | 0.180 | $8.30 \times 10^{-13}$ | -0.008 | $6.70 \times 10^{-111}$ | -0.811 | $1.01 \times 10^{-16}$ |
| 15 | rs2286528 | 17 | 59471372 | ncRNA_intronic | BCAS3:RP11-332H18.5 | T/C | 0.055 | -1.804 | 0.289 | $4.26 \times 10^{-10}$ | -0.007 | $1.21 \times 10^{-29}$ | -1.090 | $3.56 \times 10^{-12}$ |
| 16 | rs112510641 | 19 | 38254598 | intronic | ZNF573 | A/G | 0.434 | 0.733 | 0.130 | $1.74 \times 10^{-08}$ | 0.003 | $3.44 \times 10^{-32}$ | 0.420 | $2.52 \times 10^{-09}$ |

EA=effect allele, NEA = non-effect allele, EAF = effect allele frequency, SE=standard error.

Sensitivity analyses using CrCl unadjusted for BSA, with and without height as a covariate, yielded consistent results (Supplementary Table S9, Supplementary Figure S8).

Additionally, 65 loci (Supplementary Table S10) with 95 lead SNPs (Supplementary Table S11) were significantly associated with eGFR_crea_ (Supplementary Figure S9). All eGFR_crea_ loci found in Lifelines (Supplementary Table S10) overlapped with previously loci reported in CKDGen, except rs7524078 (chr 1, locus coordinates: 201992736-202015940). Interestingly, the top SNPs for eGFRcrea on chromosome 15, were not significant for CrCl (Figure 1, Supplementary Table S12). For example, rs1346267 (Supplementary Figure S10-S11) showed the strongest association with eGFRcrea (*P* = 1.53 × 10⁻⁶¹), but not for CrCl (*P* = 0.0003).

### Sex-specific GWAS meta-analysis

We identified 9 loci with 11 lead SNPs significantly associated with CrCl in females; and 2 loci with 3 lead SNPs in males (Figure 2; Supplementary Table S13-S14). One new locus in females on chromosome 13 (rs8002366, *P* = 8.44x10^-9^) had not been previously linked to eGFR.

**Figure 2.**
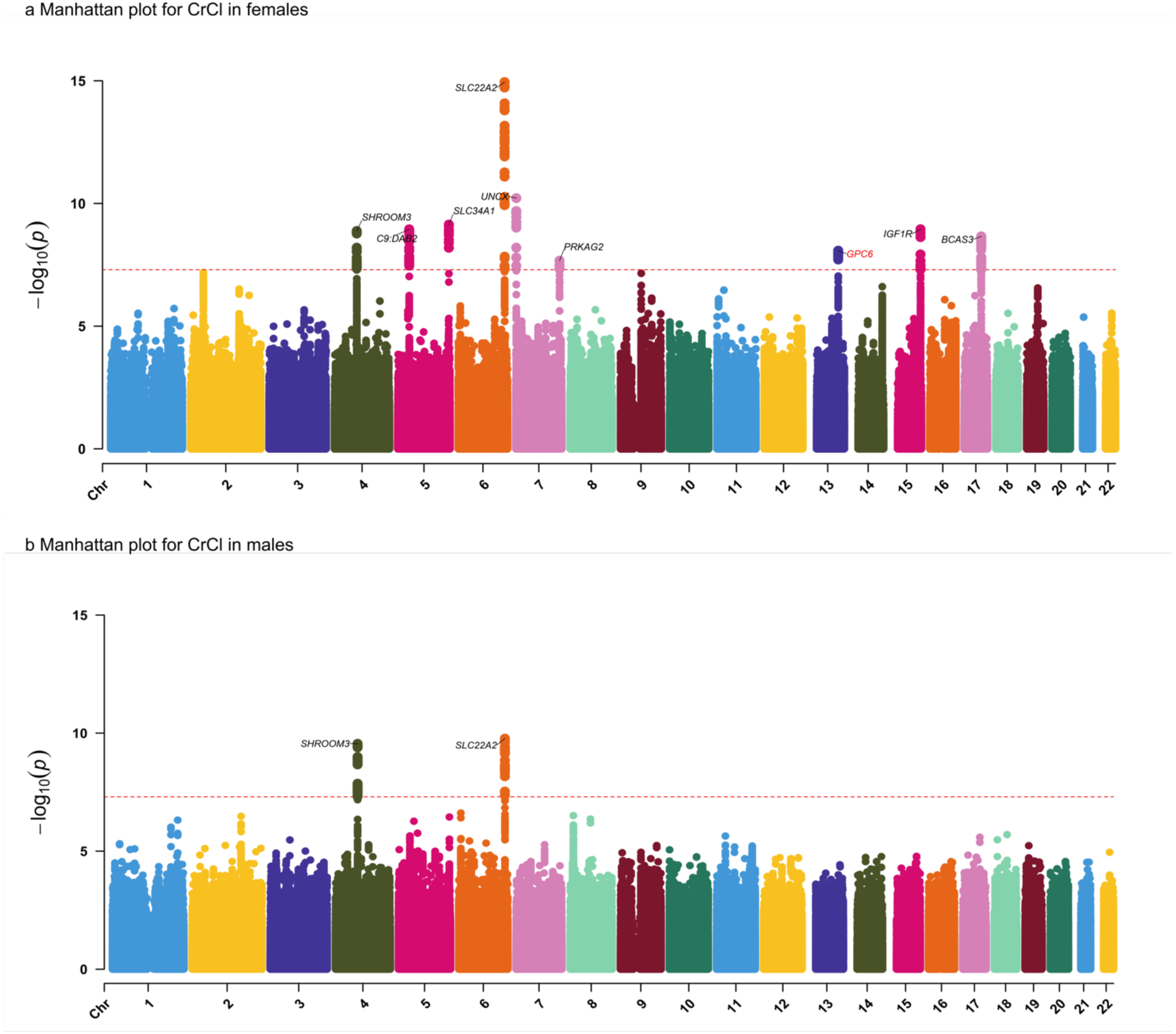
Manhattan plots of sex-specific GWAS results. The Manhattan plots show –log10 association P values for the genetic effects on creatinine clearance in females (2a) and males (2b) by chromosomal base position (GRCh37). The red line indicates the threshold for genome-wide significance (*P* < 5 × 10⁻⁸). Models were adjusted for age and 20 principal components (PCs). *GPC6,* highlighted in red, represents novel signal associated with kidney function. *IGF1R* was identified as a female-specific signal and has been previously reported to be associated with kidney function in sex-combined GWAS (2a). Panel titles: 2a Manhattan Plot for CrCl in Females, 2b. Manhattan Plot for CrCl in Males

We compared lead SNPs across sexes and with the combined model. Eight of the eleven female-associated SNPs were not significant in males; six were significant in the combined model, while two SNPs were female-specific (rs8002366, *P* = 8.44 × 10^-9^; rs12908437, *P* = 1.12 × 10^-9^) (Table 2).

**Table 2.** Sex-specific GWAS results for CrCl.

| CrCI |  |  |  |  |  |  | Females |  |  | Males |  |  | Sex-combined model |  |  | Unique |  |
| --- | --- | --- | --- | --- | --- | --- | --- | --- | --- | --- | --- | --- | --- | --- | --- | --- | --- |
| GWAS Model | Locus | SNP | Chr | Position | Type | Nearest Gene | Effect | SE | Pvalue | Effect | SE | Pvalue | Effect | SE | Pvalue | Sex-Diff Pvalue* | sex-specific SNP |
| F | 1 | rs7676094 | 4 | 77412894 | intronic | SHROOM3 | -0.967 | 0.159 | $1.33 \times 10^{-9}$ | -1.296 | 0.213 | $1.15 \times 10^{-9}$ | -1.073 | 0.130 | $1.36 \times 10^{-16}$ | 0.044 | no |
| F | 2 | rs35969577 | 5 | 39401384 | intronic | C9:DAB2 | -0.978 | 0.161 | $1.14 \times 10^{-9}$ | -0.915 | 0.213 | $1.73 \times 10^{-5}$ | -0.963 | 0.130 | $1.54 \times 10^{-13}$ | 0.677 | no |
| F | 3 | rs6862195 | 5 | 176822512 | intronic | SLC34A1 | -1.034 | 0.168 | $7.32 \times 10^{-10}$ | -0.902 | 0.226 | $6.64 \times 10^{-5}$ | -0.993 | 0.137 | $4.39 \times 10^{-13}$ | 0.413 | no |
| F | 4 | rs2279463 | 6 | 160668389 | intronic | SLC22A2 | 1.822 | 0.243 | $6.81 \times 10^{-14}$ | 2.018 | 0.323 | $4.04 \times 10^{-10}$ | 1.892 | 0.198 | $1.04 \times 10^{-21}$ | 0.395 | no |
| F | 4 | rs316035 | 6 | 160696544 | ncRNA_intronic | SLC22A2 | 1.736 | 0.217 | $1.13 \times 10^{-15}$ | 1.785 | 0.289 | $6.05 \times 10^{-10}$ | 1.768 | 0.176 | $1.21 \times 10^{-23}$ | 0.809 | no |
| F | 5 | rs10277115 | 7 | 1285195 | intergenic | UNCX | 1.252 | 0.191 | $6.06 \times 10^{-11}$ | 0.330 | 0.253 | 0.1926 | 0.906 | 0.155 | $5.34 \times 10^{-9}$ | 0.0002 | no |
| F | 6 | rs10480299 | 7 | 151405818 | intronic | PRKAG2 | 1.017 | 0.182 | $2.16 \times 10^{-8}$ | 0.827 | 0.243 | 0.0007 | 0.972 | 0.148 | $5.07 \times 10^{-11}$ | 0.278 | no |
| **F** | **7** | **rs8002366** | **13** | **93999944** | **intronic** | **GPC6** | **-1.768** | **0.307** | **$8.44 \times 10^{-9}$** | **0.556** | **0.414** | **0.1787** | **-0.864** | **0.251** | **0.0006** | **0.000003** | **yes** |
| **F** | **8** | **rs12908437** | **15** | **99287375** | **intronic** | **IGF1R** | **-1.003** | **0.165** | **$1.12 \times 10^{-9}$** | **-0.362** | **0.221** | **0.1008** | **-0.703** | **0.134** | **$1.69 \times 10^{-07}$** | **0.001** | **yes** |
| F | 9 | rs6504021 | 17 | 59240473 | intronic | BCAS3 | -1.154 | 0.193 | $2.20 \times 10^{-9}$ | -0.583 | 0.257 | 0.0232 | -0.972 | 0.157 | $6.18 \times 10^{-10}$ | 0.007 | no |
| F | 9 | rs11657044 | 17 | 59450105 | ncRNA_intronic | BCAS3 | -1.283 | 0.222 | $6.84 \times 10^{-9}$ | -1.178 | 0.295 | $6.35 \times 10^{-5}$ | -1.290 | 0.180 | $8.30 \times 10^{-13}$ | 0.612 | no |
| M | 1 | rs10008637 | 4 | 77414144 | intronic | SHROOM3 | 0.900 | 0.159 | $1.63 \times 10^{-8}$ | 1.346 | 0.213 | $2.80 \times 10^{-10}$ | 1.043 | 0.130 | $9.35 \times 10^{-16}$ | 0.010 | no |
| M | 2 | rs316020 | 6 | 160669081 | intronic | SLC22A2 | 1.808 | 0.234 | $1.22 \times 10^{-14}$ | 1.952 | 0.314 | $5.13 \times 10^{-10}$ | 1.896 | 0.191 | $3.64 \times 10^{-23}$ | 0.519 | no |
| M | 2 | rs3127572 | 6 | 160682218 | intronic | SLC22A2 | 1.563 | 0.239 | $5.72 \times 10^{-11}$ | 2.031 | 0.318 | $1.69 \times 10^{-10}$ | 1.747 | 0.194 | $2.31 \times 10^{-19}$ | 0.054 | no |

CrCI GWAS Model = Indicates the source model (sex-specific analysis) from which the lead SNPs were identified: F = Females, N = 35,621; M = Males, N = 23,355. \*P value assessing sex difference based on t-statistics. Bonferroni threshold < 0.004.

Sex-difference tests across the 14 lead SNPs (11 female, 3 male) identified significant differences for rs8002366 (*P* = 0.000003) and rs12908437 (*P* = 0.001), highlighting two female-specific loci (Table 2, Supplementary Figure S12). All three male-associated SNPs were also significant in females and the combined model. Results were consistent in sensitivity analyses (Supplementary Table S15).

### Comparison with eGFR GWASs

Effect directions of the 21 lead CrCl SNPs were generally consistent with CKDGen results of both eGFRcrea and eGFRcys (Supplementary Figure S13). All lead CrCl SNPs were genome-wide significant for eGFRcrea in CKDGen, except the two novel SNPs: rs146465192 (*P* = 0.01) and rs117014836 (*P* = 0.002) (Table 1, Supplementary Table S16). Both were (suggestively) associated with eGFRcrea in Lifelines (rs146465192: *P* = 1.34 × 10^-8^; rs117014836: *P* = 3.64 × 10^-7^) (Table 1, Supplementary Table S17). For Cystatin C (eGFR_cys_) in CKDGen, rs146465192 was unavailable, and rs117014836 was not associated (*P* = 0.566) (Supplementary Table S18).

Additionally, we compared the 424 lead SNPs reported in CKDGen for eGFRcrea and the 323 lead SNPs associated with eGFRcys with our CrCl GWAS results in Lifelines (Supplementary Table S19). Overall, effect directions were largely consistent (Supplementary Figures S14-S15). Interestingly, the strongest SNP for eGFRcrea in CKDGen (*SPATA5L1,* rs2433601, *P* = 1.13 × 10⁻²⁶²), was not genome-wide significant for CrCl (*P* = 0.0004) (Supplementary Figure S14), but overlaps with the strongest locus associated with eGFR in Lifelines (15:45690989:C:T; rs1346267; *P* = 1.53 × 10⁻⁶¹).

We also compared sex-specific CrCl lead SNPs with eGFR GWAS data. One female-specific SNP (rs12908437) was replicated for (sex-combined) eGFR_crea_ and eGFR_cys_ in CKDGen (Supplementary Table S20). In Lifelines, neither of the two female-specific CrCl loci was genome-wide significant in the sex-combined eGFR_crea_ GWAS (rs8002366, *P* = 0.0007; rs12908437, *P* = 4.5 × 10^-4^) (Supplementary Table S21). Furthermore, they were also sex-specific for eGFR_crea_ in Lifelines (Supplementary Table S22). rs8002366 passed the Bonferroni correction for sex differences, while rs12908437 showed nominal significance (*P* < 0.05).

### Shared genetic architecture and SNP-based heritability

CrCl showed high genetic correlation with eGFR_crea_ from CKDGen (rg = 0.73, *P* = 2.36 × 10^-89^) and from Lifelines (rg = 0.77, *P* = 8.50 × 10^-124^), whereas the genetic correlation with eGFRcys was more moderate (rg = 0.48, *P* = 1.41 × 10^-20^) (Supplementary Table S23). SNP-based heritability was 12% for CrCl.

Sex-specific CrCl analyses showed genetic correlations of 0.72 (*P* = 1.41 × 10⁻⁴⁹) in females and 0.70 (*P* = 1.37 × 10⁻³⁰) in males with sex-combined eGFRcrea from CKDGen. These correlations were higher with sex-specific eGFR_crea_ from Lifelines: 0.78 (*P* = 3.30 × 10⁻⁵⁸) in females and 0.82 (*P* = 1.55 × 10⁻⁴⁰) in males. Corresponding h² estimates were 12% in females and 14% in males. Genetic correlations with eGFRcys are provided in Supplementary Table S23.

### Bioinformatics follow-up

In silico lookups showed that 19 of the 21 SNPs had been previously associated with kidney function phenotypes. For the two new SNPs, no linked variants or reported associations were found for rs146465192. For rs117014836, three linked intronic variants (*MAP3K4*-rs183271809, *AGPAT4*-rs117062161, and *AGPAT4*-rs187184205) were identified, but none had reported associations (Supplementary Table S24).

### eQTL, pQTL and PheWAS lookup

rs117014836 was associated with *AGPAT4* expression in blood (eQTL *P* = 6.54 × 10^-6^) (Supplementary Table S25). pQTL lookups revealed that the A allele of rs146465192 was negatively associated with plasma *IGF2R* (β = -0.519, *P* = 1.40 × 10^-6^). PheWAS also showed this allele associated with low blood density lipoprotein (LDL) cholesterol (β = 0.044, *P* = 1.80 × 10^-6^).

eQTL lookups of our two sex-specific SNPs revealed rs12908437 associated with *IGF1R* expression in blood (*P* = 2.62 × 10^-6^), and rs8002366 with *GPC6* expression in kidney (*P* = 8.38 × 10^-10^) (Supplementary Table S25).

### Prioritization of kidney cell types

Cell type prioritization for CrCl in mature human kidney identified five cell types with nominal significance (*P* < 0.05; Figure 3): proliferating proximal tubule, proximal tubule, connecting tubule, epithelial progenitor cells, and distinct proximal tubule 2 (Figure 3). Mean expression of *IGF2R*, *AGPAT4*, and female-specific loci *IGF1R* and *GPC6*, align with tubular epithelial cell prioritization (Table 3, Supplementary Figures S16-S17).

**Figure 3.**
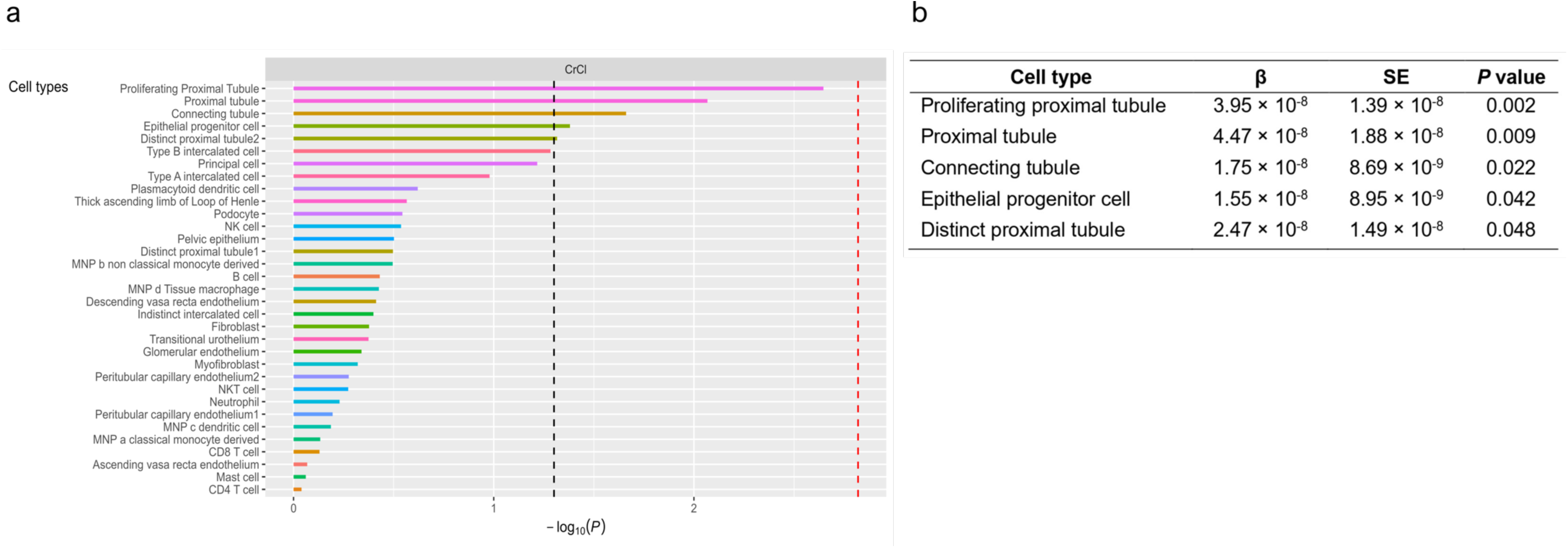
Cell type prioritization within the mature human kidney was estimated using LDSC across CrCl GWAS data. The results display the association strength as -log10 (*P* value) between cell type expression specificity and CrCl heritability. Cell types are sorted in descending order of -log10 *P* values across the GWAS data. The dashed black line indicates the nominal significance threshold (*P* < 0.05), while the red dashed line marks the Bonferroni significance threshold (0.05/33, *P* = 0.002) (3a). The table displays the Beta (β), standard error (SE), and P values for the LDSC integration of CrCl GWAS data with cell types from mature human kidney tissue (3b).

**Table 3.** Phenotypic impact of prioritized CrCl SNPs.

| GWAS model | SNP | Chr:Position | Novel<br>Kinney<br>Function<br>SNP | GWAS catalog known<br>associations | eQTL | pQTL | PheWAS | snRNA predominant<br>kidney cell type* |
| --- | --- | --- | --- | --- | --- | --- | --- | --- |
| Sex-combined | rs146465192 | 6:160297118 | yes | NF | NF | *IGF2R*,<br>$\beta = -0.519$,<br>$P = 1.40 \times 10^{-6}$ | Plasma LDL,<br>$\beta = 0.044$,<br>$P = 1.80 \times 10^{-6}$ | Plasmacytoid dendritic cell<br>Proximal tubular cell<br>NK cell<br>Myofibroblast<br>Epithelial progenitor cell |
| Sex-combined | rs117014836 | 6:161516388 | yes | NF | *AGPAT4*,<br>Gene in blood<br>$P = 6.54 \times 10^{-6}$ | NF | NF | NK cell<br>Peritubular capillary endothelium<br>Glomerular endothelium<br>Proximal tubular cell<br>Connecting tubule |
| Females | rs8002366 | 13:93999944 | yes | NF | *GPC6*,<br>Gene in kidney<br>$\beta = -0.284$,<br>$P = 8.38 \times 10^{-10}$ | NF | NF | Fibroblast<br>Podocyte<br>Myofibroblast<br>Proximal tubular cell<br>Epithelial progenitor cell |
| Females | rs12908437 | 15:99287375 | no | Atrial fibrillation, Blood<br>urea nitrogen levels,<br>Blood urea nitrogen<br>levels, eGFR, Serum<br>creatinine levels, Serum<br>uric acid levels | *IGF1R*,<br>Gene in blood,<br>$P = 2.62 \times 10^{-6}$ | NF | NF | Proximal tubular cell<br>Urothelial cell<br>Epithelial progenitor cell<br>Podocyte<br>Peritubular capillary endothelium |

SNP = single nucleotide polymorphisms; eQTL = expression quantitative trait loci; pQTL= protein quantitative trait loci; PheWAS = Phenome-wide association studies; NF =
Not found in post-GWAS lookups; \*Top five cell types with highest gene expression (Supplementary Figures S16-S17). NK cell: natural killer cell.

### Colocalization and fine-mapping analyses

Colocalization analyses of the 2 novel and 2 sex-specific loci did not provide evidence for shared causal variants between GWAS loci and tested eQTL or pQTL signals (Supplementary Figures S18-S19, Supplementary Table S26-S27, Supplementary Results). Fine-mapping across these four CrCl loci, highlighted a small set of candidate variants with high posterior probabilities, supporting them as strong candidates for functional follow-up (Supplementary Figures S20-S21, Supplementary Table S28-S29, Supplementary Results).

## Discussion

In this first large-scale GWAS of CrCl in 58976 Europeans, we identified 16 loci with 21 independent genome-wide significant lead SNPs, including two novel SNPs on chromosome 6 (rs117014836 and rs146465192). Functional analyses linked rs117014836 to expression levels of Acylglycerophosphate Acyltransferase 4 (*AGPAT4*) in blood, and rs146465192 to plasma levels of insulin-like growth factor 2 receptor (*IGF2R*) and LDL cholesterol. Sex-stratified analyses identified two female-specific loci: Insulin Like Growth Factor 1 Receptor (*IGF1R*, rs12908437) and Glypican 6 (*GPC6*, rs8002366), with rs8002366 as a novel locus. These findings enhance understanding of genetic determinants of CrCl, a muscle-mass independent kidney function measure.

*AGPAT4* encodes an isoform of the acylglycerophosphate acyltransferase/lysophosphatidic acid acyltransferase (*AGPAT/LPAAT*) family, catalyzing lysophosphatidic acid conversion to phosphatidic acid^48,49^. These enzymes transfer acyl groups to lysophospholipids, generating complex phospholipids^50^. A recent study combining metabolomics in diabetic kidney disease (DKD) patients, an animal model, and *in vitro* experiments demonstrated that disruptions in lysophospholipid metabolism contribute to kidney function decline^51^. Increased urinary lysophosphatidylcholine (LPC) was linked to faster kidney function decline validated in animals showing that lipid metabolism imbalance leads to abnormal LPC accumulation in the kidney^51^. In vitro, LPC impaired autophagic flux and increased stress in renal tubular cells, a key pathogenic mechanism in DKD progression^51^. Research on AGPAT4 is limited; previous studies reported associations with lipid metabolism^52^, cancer^53,54^, cognitive impairment^55^, and skeletal muscle function and fiber composition^56^. Importantly, rs117014836 had not been reported in any previous kidney function GWAS.

The second SNP rs146465192, also on chromosome 6, overlaps with a previously reported eGFR-associated locus in CKDGen (rs3119304, nearest gene *SLC22A2*). These variants are not in strong LD (r^2^ = 0.050, 1000 Genomes Project, CEU)^46,47^, indicating independent associations. Functional follow-up revealed rs146465192 is associated with plasma *IGF2R* levels, suggesting a potential regulatory role*. IGF2R* is a multifunctional receptor modulating IGF2, a key growth factor regulating cell proliferation, apoptosis, and metabolism^57,58^. IGFs (IGF-1 and IGF-2) are essential for kidney development and function, influencing renal hemodynamics both directly and indirectly through interactions with the renin-angiotensin system^59,60^. Disruptions in IGF signaling have been linked to CKD, diabetic nephropathy, polycystic kidney disease, and renal fibrosis^58,59,61^. Additionally, rs146465192 is associated with LDL cholesterol levels, reinforcing the connection between lipid metabolism and kidney function.

In addition to the two novel CrCl variants among both sexes, we identified two female-specific loci associated with lower CrCl. *IGF1R* (rs12908437) supports the role of IGF signaling in kidney function. *IGF1R* encodes a tyrosine kinase receptor activated by IGF-1, IGF-2, and insulin, playing a key role in cell growth and proliferation^62^. Although previous GWAS linked *IGF1R* to kidney function, these were based on sex-combined analyses^3^. Importantly, sex differences in *IGF1R* have been reported in metabolic^63^ and hormonal regulation^64^. One study identified G310D, a missense variant in *IGF1R* on chromosome 15 (distinct from our variant; not in public LD reference panels), as associated with increased risk of type 2 diabetes in females^65^.

Additionally, females carrying the *GPC6* rs8002366-T allele had a lower CrCl, and this allele was associated with reduced *GPC6* expression in the kidney. *GPC6* encodes glypican-6, a heparan sulfate proteoglycan involved in signaling pathways regulating cell growth, differentiation, and development, such as Wnt, Hedgehog, and fibroblast growth factors^66,67^. Proteoglycans contribute to kidney function by maintaining the glomerular filtration barrier and regulating cytokines^68–71^. While no previous studies have directly linked GPC6 to kidney function, other glypicans have been implicated in kidney diseases: *GPC1* in fluid balance regulation and renal response^72^, *GPC3* in Wilms tumors and renal malformations^73–75^, *GPC4* in low kidney function^75,76^, and *GPC5* in nephrotic syndrome^77,78^. Further research is needed to clarify the role of GPC6 in kidney function and its sex-specific effects.

Two previous GWASs on kidney function reported mixed findings regarding sex differences; one found none^79^, while the other identified a single female-specific locus^80^. Previous research has shown sex differences in kidney function and related diseases^9–12^, involving mechanisms such as hormonal regulation^81,82^, renal transport^83,84^, age-related kidney function decline^13^, sexual dimorphism in kidney morphology and function^85^, and differential disease susceptibility^9–12^.

Consistent with previous studies^3^, our cell type prioritization analysis highlighted proximal tubular epithelial and epithelial progenitor cells as key contributors to kidney function. This is further supported by the observation that all four prioritized genes are also predominantly expressed by tubular epithelial cells (Figure 3). Together, these results provide complementary evidence linking genetic associations with functional cell types that influence kidney function.

Although colocalization analyses did not support shared causal variants between CrCl GWAS loci and the tested QTLs, the consistent identification of strong and independent signals in the same genomic regions is noteworthy. The fact that lead GWAS variants coincide with robust cis-eQTL and pQTL associations, underscores the biological relevance of these loci and highlights them as promising candidates for functional follow-up.

This is, to our knowledge, the first GWAS investigating genetic variants influencing CrCl as a muscle mass-independent phenotype. When comparing our findings with eGFR_crea_ GWASes, our two sex-combined SNPs were suggestively associated with eGFR_crea_ in Lifelines, but not in CKDGen. One was not found in the eGFR_cys_ dataset from CKDGen, and the second was not associated. The female-specific SNP (rs12908437) was replicated for sex-combined eGFR_crea_ and eGFR_cys_ in CKDGen.

Notably, our strongest lead SNP for eGFR_crea_ (rs1346267, *P* = 1.53 × 10⁻⁶¹) was not associated with CrCl (*P*=0.0003). This locus overlaps with the *SPATA5L1* locus reported by CKDGen (rs2433601; eGFR_crea_ *P*=1.13×10^-262^, eGFR_cys_ *P*=0.790)^3^ and shows high LD (r² = 0.99)^86^. Both rs1346267 and rs2433601 have been linked to *GATM* splicing^87,88^, a gene involved in creatinine metabolism^89,90^. The lack of CrCl association supports the interpretation that this locus influences creatinine production, rather than glomerular filtration itself. This example highlights a genetic signal that affects creatinine without necessarily reflecting underlying kidney function.

Sensitivity analyses using CrCl unadjusted for BSA yielded similar results across sex-combined and sex-stratified GWASs, supporting the robustness of our findings. Interestingly, two loci (comprising three lead SNPs) from the unadjusted CrCl GWASs were not detected in the primary GWAS (Supplementary Figure S8). The nearest genes were *RPGRIPL*, *FTO,* and *MC4R* (Supplementary Figure S22), known obesity-related genes^91–93^. Functional annotation showed these SNPs have previously been associated with anthropometric traits (Supplementary Table S30). These SNPs were not associated with eGFR_crea_ in either Lifelines or CKDGen (Supplementary Table S31), suggesting that their association with CrCl may be driven by variation in body size or composition, rather than kidney function.

While this study provides valuable insights into the genetic architecture of CrCl, certain limitations should be acknowledged. A key strength is the use of CrCl as a muscle mass-independent measure of kidney function, providing a unique perspective compared to previous GWASs relying on eGFR_crea_. However, creatinine is not only filtered by the glomerulus but also actively secreted by renal tubules, which can lead to an overestimation of kidney function, especially in individuals with impaired kidney function. Our analyses were conducted in individuals of European ancestry, limiting generalizability. Additional studies in diverse populations are needed to validate these findings and to explore sex-specific differences, improving understanding of underlying biological mechanisms. At the same time, the scarcity of 24-hour urine collections in large cohorts limits possibilities for external replication.

In conclusion, the present study identified two novel signals implicated in kidney function through evaluation of creatinine clearance (*AGPAT4*-rs117014836 and *IGF2R*-rs146465192). Additionally, we identified two unique sex-specific loci, one of which was novel (*GPC6*-rs8002366). These findings provide new insights into the genetic architecture of kidney function and highlight the value of CrCl as a distinct kidney function phenotype for genetic studies.

## Data Availability

Data may be obtained from Lifelines through a controlled access data repository. More information about how to request Lifelines data and the conditions of use can be found on their website (https://www.lifelines-biobank.com/researchers/working-with-us). Full GWAS summary statistics of our meta-analyses will be publicly available on the GWAS Catalog website data repository (https://www.ebi.ac.uk/gwas/).

## Acknowledgements

The authors wish to acknowledge the services of the Lifelines Cohort Study, the contributing research centers delivering data to Lifelines, and all the study participants.

## Author contributions

A.D.A.P., P.J.v.d.M., H.S., and M.H.d.B. wrote the first draft of the manuscript; P.J.v.d.M., H.S., and M.H.d.B. conceived and designed the project; A.D.A.P., P.J.v.d.M., A.V., S.J.L.B., H.S., and M.H.d.B. contributed to methodology development; A.D.A.P. performed the association analyses, meta-analyses and follow-up analyses; Z.K. performed the eQTL, pQTL, and PheWAS lookups; Z.K. and M.G.A. conducted the cell type prioritization, colocalization and fine-mapping analyses; All authors revised and edited the manuscript; H.S. and M.H.d.B. supervised and led the project.

