## Supplementary Figure for "Genome-Wide Association Study of Creatinine Clearance Identifies New Loci for Kidney Function"

### Supplementary material

Supplementary Figure S19. Colocalization results for the two female-specific CrCl-associated SNPs 27

Supplementary Figure S20. Fine-mapping results for the two novel CrCl-associated SNPs................28

Supplementary Figure S21. Fine-mapping results for the two female-specific CrCl-associated SNPs 29

Supplementary Figure S22. Regional plots for three lead SNPs exclusive to CrCl sensitivity GWAS models. ..........................................................................................................................................30

**Supplementary Note**

**UMCG Genetics Lifelines Initiative (UGLI) group author: LifeLines Cohort Study**

Raul Aguirre-Gamboa[1], Patrick Deelen[1], Lude Franke[1], Jan A Kuivenhoven[2], Esteban A Lopera Maya[1], Ilja M Nolte[3], Serena Sanna[1], Harold Snieder[3], Morris A Swertz[1], Peter M. Visscher[3,4], Judith M Vonk[3], Cisca Wijmenga[1], Naomi Wray[4].

[1] Department of Genetics, University of Groningen, University Medical Center Groningen, The Netherlands

[2] Department of Pediatrics, University of Groningen, University Medical Center Groningen, The Netherlands

[3] Department of Epidemiology, University of Groningen, University Medical Center Groningen, The Netherlands

[4] Institute for Molecular Bioscience, The University of Queensland, Brisbane, Queensland, Australia.

#### Supplementary Methods, Results, Discussion and References

##### Distributional assumptions

We assessed the distribution of CrCl and eGFR visually using histograms in the total sample and stratified by cohort. CrCl showed an approximately normal and symmetric distribution across cohorts, and therefore no transformation was applied. Although eGFR showed a mild deviation from normality, given the large sample size and the robustness of linear regression to moderate departures from normality[S1] we retained eGFR on its original scale to preserve interpretability.

##### Prioritization of kidney cell types for creatinine clearance: Supplementary Methods

- **Single cell RNA-seq dataset**

We used data from Stewart et al., which includes 33 cell type annotations from the cortex and medulla of 14 mature human kidneys.[40]

- **GWAS data pre-processing**

We adapted GWAS summary statistics of creatinine clearance (CrCl) indices in Europeans using the 'munge_sumstats.py' script from the LDSC v1.0.0 package for cell type prioritization.[33] The datasets were restricted to HapMap3 SNPs, excluding those within the major histocompatibility complex region (chr6:25Mb-34Mb).

- **Single-cell RNA-seq data pre-processing**

The single-cell RNA-seq dataset was annotated to cell types based on the original publication and processed uniformly using the CELLEX (CELL type Expression-Specificity) toolkit.[42] The gene expression matrix was normalized and log-transformed, followed by computation of the expression specificity matrix. Expression Specificity Weights (ESw) were derived using four metrics, and average expression specificity scores (ESμ) were calculated for each gene within cell types. Cell type prioritization was then based on ESμ estimates.[42]

- **Cell type prioritization based on LD score regression analysis**

We analyzed cell type prioritization using the CELLECT (CELL type Expression-specific integration for Complex Traits) toolkit[42] combined with the stratified linkage disequilibrium score (S-LDSC) algorithm. This approach was designed to assess whether specific cell types, identified by average specificity metrics, were enriched in heritability for the CrCl trait. A 100-kilobase window around genes' transcribed regions was used to capture regulatory elements influencing trait effects. Annotation files were created for each cell type, assigning gene ESμ values to variants within the 100-kb window. LD scores were calculated for HapMap3 SNPs, and S-LDSC was applied to generate one-tailed p-values for positive associations between trait heritability and cell type ESμ values. Regression effect sizes for each cell type were estimated to represent the per-SNP heritability change due to increased expression specificity, with standard errors calculated using the jackknife method.[42] The significance level was determined based on the Bonferroni p-value for the kidney tissue dataset, alongside the nominal p-value.

##### Fine-mapping analyses: Supplementary Methods

We performed fine-mapping analysis of creatinine clearance GWAS loci using FINEMAP v1.4.2.[45] We started with novel index SNPs and expanded each locus to include all variants within ±500 kb. For each region, we constructed z-files containing SNP-level summary statistics and generated corresponding linkage disequilibrium (LD) matrices using the European reference panel from the 1000 Genomes Project Phase,[30] computed with PLINK v1.9.[S2] FINEMAP was then run with default settings to estimate the posterior probability of causality for each SNP and to identify the most likely causal variants within each locus.

##### Fine-mapping analyses: Supplementary Results

Fine-mapping identified several candidate variants with high posterior inclusion probabilities (PIP) across the four genomic regions. At 6q25.3 (index SNP: rs146465192), multiple variants showed compelling evidence of causality, with PIP values approaching 1.0. Notably, rs316031, rs653753, rs421913, and rs316009 each had PIP = 1.0 and $\log_{10}$ Bayes factors > 13, indicating very strong support. These variants were also associated with large effect sizes (e.g., rs316009: $\beta = 1.89$, SE = 0.19, z = 9.88). At 6q26 (index SNP: rs117014836), the fine-mapping similarly highlighted multiple likely causal variants, including rs117189174 and rs149851855, both with PIP = 1.0 and $\log_{10}$ Bayes factors > 13.

For the female-specific locus on chromosome 13 (index SNP: rs12875395), three variants—rs12875395, rs7317246, and rs17267683—emerged as strong candidates, each with PIP = 1.0 and $\log_{10}$ Bayes factors > 12. At the other female-specific locus on chromosome 15, three variants also showed compelling evidence for causality: the index SNP rs12908437, along with rs61168554 and rs11855223.

Taken together, the fine-mapping narrowed down each of the four-creatinine clearance GWAS loci to a small set of candidate variants with high posterior probabilities, highlighting strong candidates for functional follow-up.

We used Ensembl Biomart to identify possible consequences and associations of the prioritized likely causal SNPs. At the sex-specific locus, the lead SNP rs12908437 showed a high probability of being causal, together with a small set of additional variants within the same region. Variants at this locus have previously been associated with urate levels, blood urea nitrogen (BUN), body mass index (BMI), atrial fibrillation, and male puberty timing in published GWAS.

At the novel locus indexed by rs146465192, rs316009 emerged as a likely causal variant and has been previously linked to glomerular filtration rate. In addition, rs3757020 at the novel locus rs117014836 and rs421913 at the rs146465192 locus have been reported to be associated with lipoprotein(a) levels. Together, these findings suggest that the fine-mapped variants at CrCl loci overlap with genetic signals influencing renal and cardiometabolic traits, providing biological plausibility for their role in kidney function.

#### Colocalization analyses: Supplementary Methods

To evaluate whether the same genetic variants underlie associations between creatinine clearance and gene expression or protein levels, we performed colocalization analyses using the coloc package version 5.2.3[43] and its fine-mapping extension coloc.susieR.[44]

We examined four candidate genes identified from QTL lookups (Table 3) based on the following resources: *AGPAT4 and IGF1R* from the blood eQTLgen consortium,[36] *GPC6* from the NephQTL2 kidney eQTL dataset,[S3] and *IGF2R* from the Sun et al. plasma proteome study (pQTLs).[S4] For eQTL datasets, we extracted all available cis-SNPs for each gene. For the pQTL, we included the GWAS lead SNP and all variants within ±1 Mb of it, to harmonize the genomic window with eQTL analyses.

For standard coloc analysis, we estimated posterior probabilities for the five hypotheses: H0: no association with either trait, H1: association with creatinine clearance only, H2: association with molecular signature only, H3: association with both traits, but different causal variants, and H4: association with both traits, shared causal variant.

We also applied coloc.susieR, which incorporates credible sets derived from SuSiE fine-mapping into coloc inference, enabling multiple causal variants to be assessed per locus. LD matrices for input SNP sets of each gene were calculated by PLINK1.9 software,[S2] based on 1000 Genomes reference panel of European individuals (phase 3, version 5a).[30] Default parameters were used, and analyses were restricted to SNPs present in both GWAS and molecular QTL datasets.

##### Colocalization analyses: Supplementary Results

Colocalization analyses provided no significant evidence that associations between creatinine clearance GWAS loci and molecular QTL signals were driven by the same causal variants.

In standard coloc analyses, all four tested loci demonstrated extremely strong support for H3 (posterior probability >0.99), indicating that both the creatinine clearance association and the molecular QTL signal are genuine but arise from distinct causal variants. Similarly, coloc.susieR analyses did not identify any shared credible sets between GWAS and QTL signals. Across all loci, SuSiE-based colocalization results showed H3 = 1.0 with no evidence for H4, further confirming that creatinine clearance signals and tested molecular traits are influenced by independent variants.

Taken together, these results suggest that while the examined eQTL and pQTL signals co-occur in genomic regions associated with creatinine clearance, the underlying causal variants are distinct. This indicates that, at least for these genes, creatinine clearance loci are unlikely to act through the tested blood, kidney, or plasma protein molecular traits.

##### Supplementary Discussion

Although colocalization analyses did not support shared causal variants (H4) between creatinine clearance GWAS loci and the tested QTLs, the consistent identification of strong and independent signals in the same genomic regions is noteworthy. The fact that lead GWAS variants coincide with robust cis-eQTL and pQTL associations underscores the biological relevance of these loci and highlights them as promising candidates for functional follow-up.

Importantly, lack of statistical colocalization should not be interpreted as absence of a mechanistic relationship. Coloc analyses rely on the simplifying assumption of a single causal variant per locus, and even in the SuSiE framework, the resolution is limited by sample size, LD complexity, and differences in study design.

Thus, our findings suggest that creatinine clearance loci harbor both GWAS and QTL signals in close proximity, especially for *GPC6*, but with currently available datasets, they appear to be driven by distinct causal variants. These results still add biological depth to our GWAS signals, as they point to genes and proteins that are strongly regulated in the same regions, providing plausible functional links even if not confirmed by strict colocalization. Future work with larger, tissue-specific datasets and integrative approaches may help disentangle the causal architecture and uncover shared mechanisms.

#### Supplementary Figures

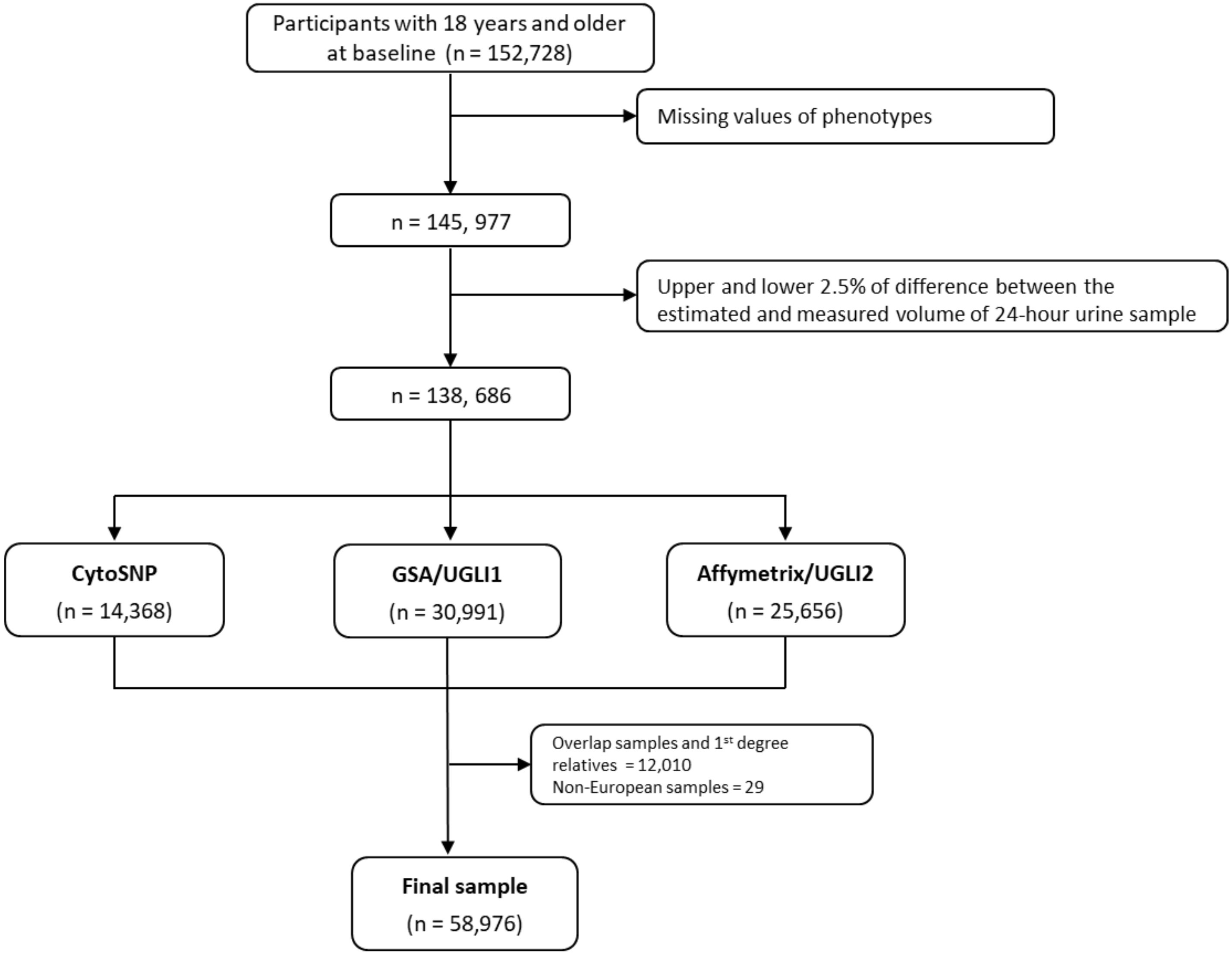

Supplementary Figure S1. Flow chart. CytoSNP, Illumina CytoSNP-12v2 array; GSA, Infinium Global Screening Array® (GSA) MultiEthnic Disease Version; Affymetrix, FinnGen Thermo Fisher Axiom® custom array

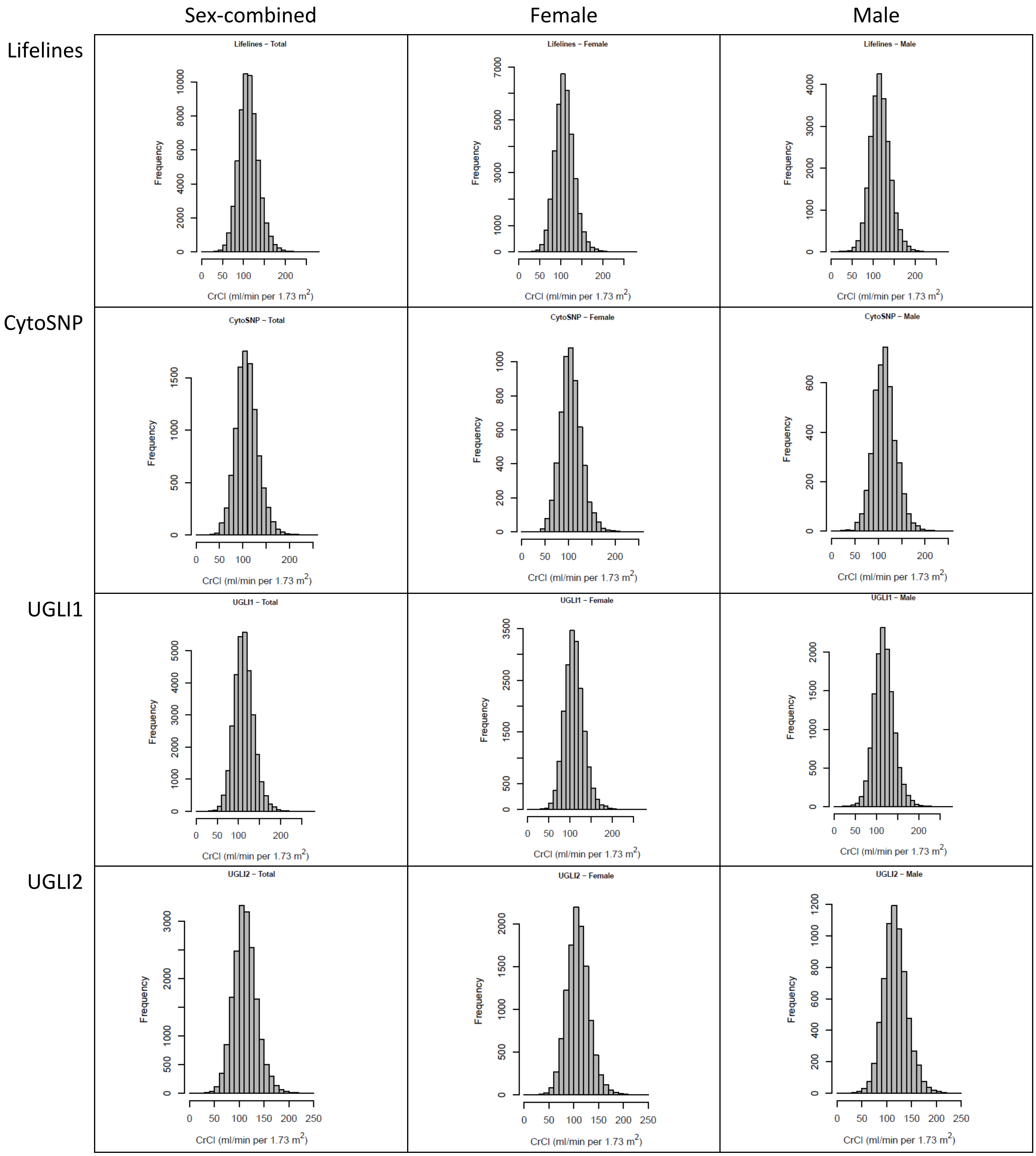

Supplementary Figure S2. Creatinine clearance distribution in sex-combined and sex-specific data, across all sample size and per subcohort.

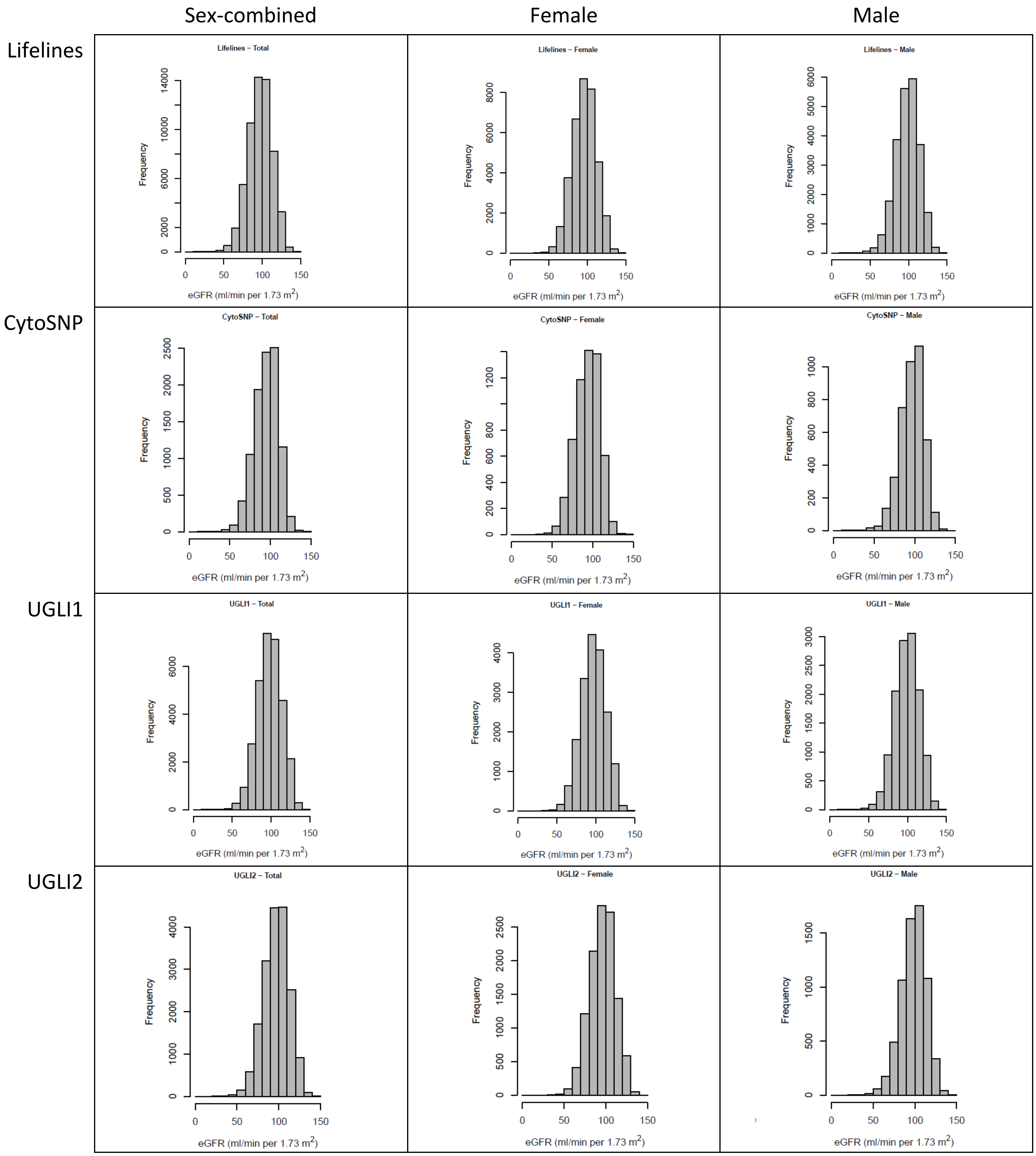

Supplementary Figure S3. eGFR distribution in sex-combined and sex-specific data, across all sample size and per subcohort.

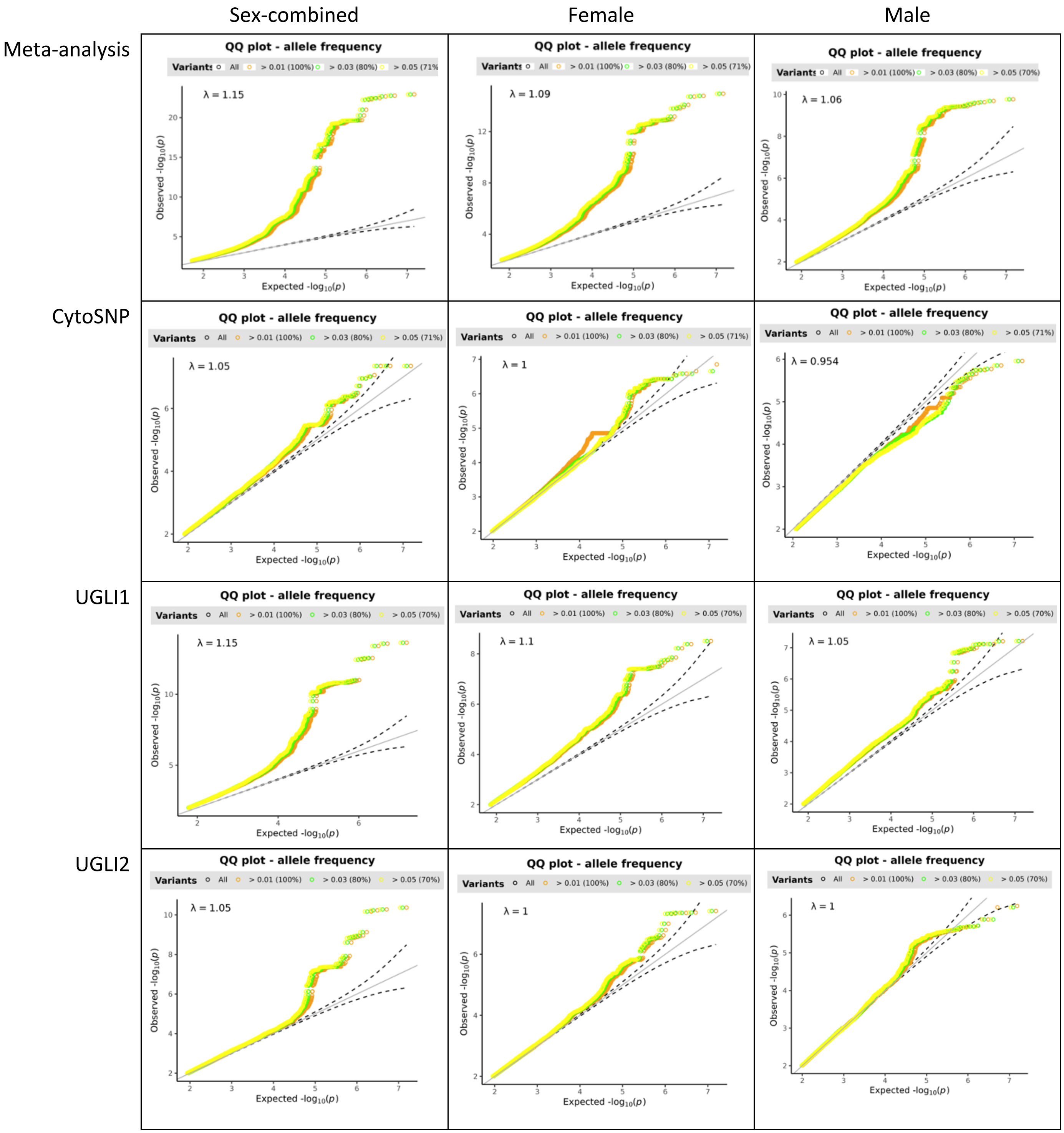

Supplementary Figure S4. Quantile-Quantile plots of the SNPs in the genome-wide association study of creatinine clearance.

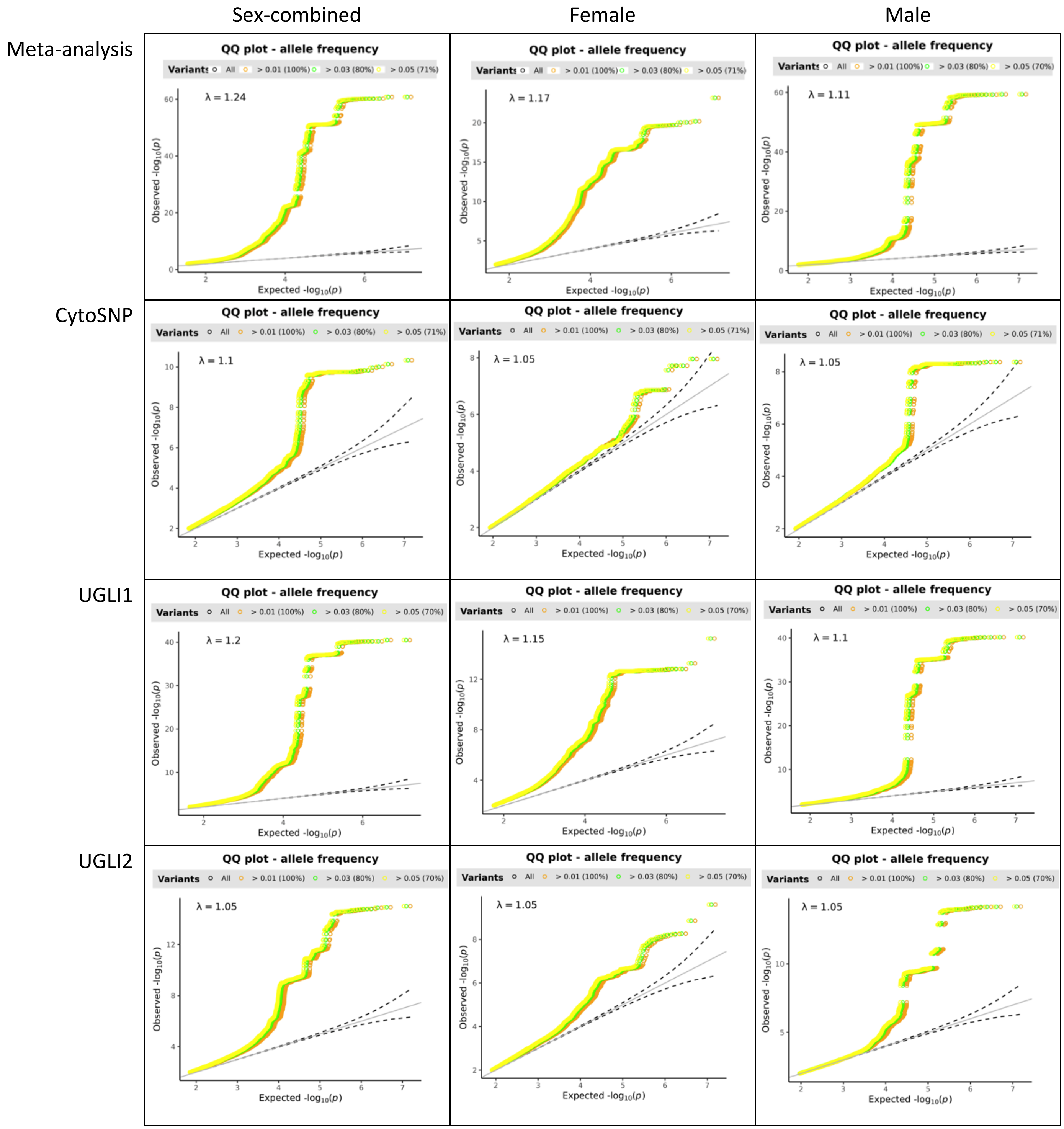

Supplementary Figure S5. Quantile-Quantile plots of the SNPs in the genome-wide association study of eGFR.

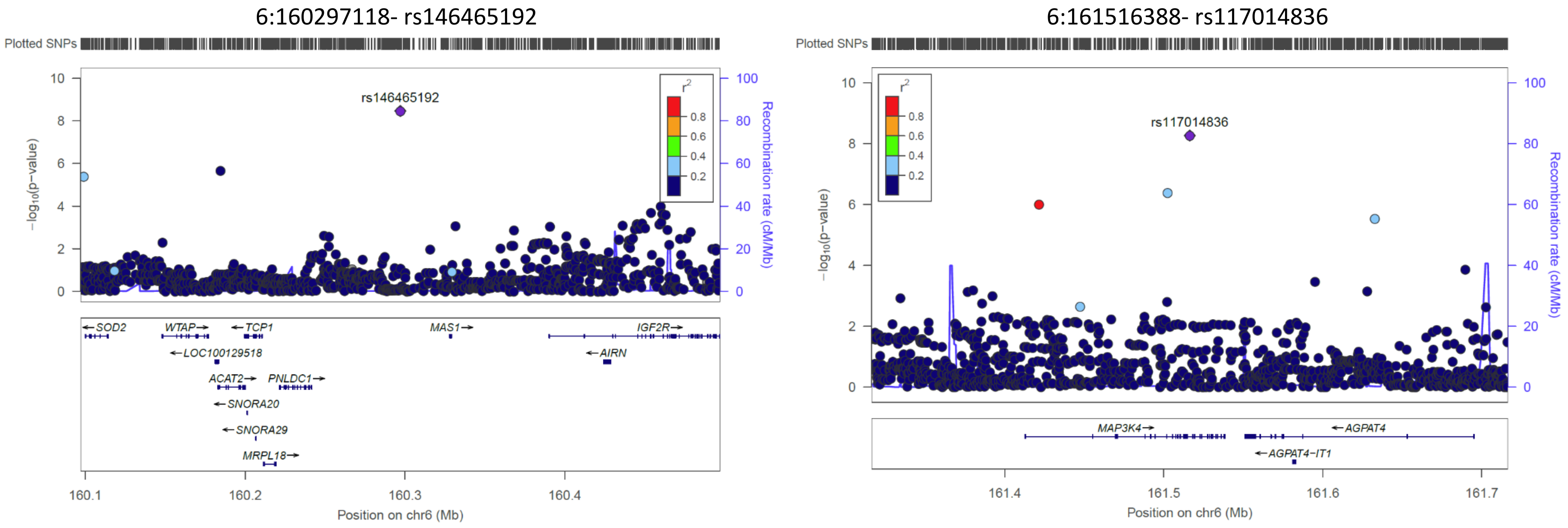

Supplementary Figure S6. Regional plots for rs146465192 and rs117014836. $r^2$ values are from hg19 / 1000 Genomes EUR reference samples. Plots were generated with LocusZoom (http://csg.sph.umich.edu/locuszoom/).

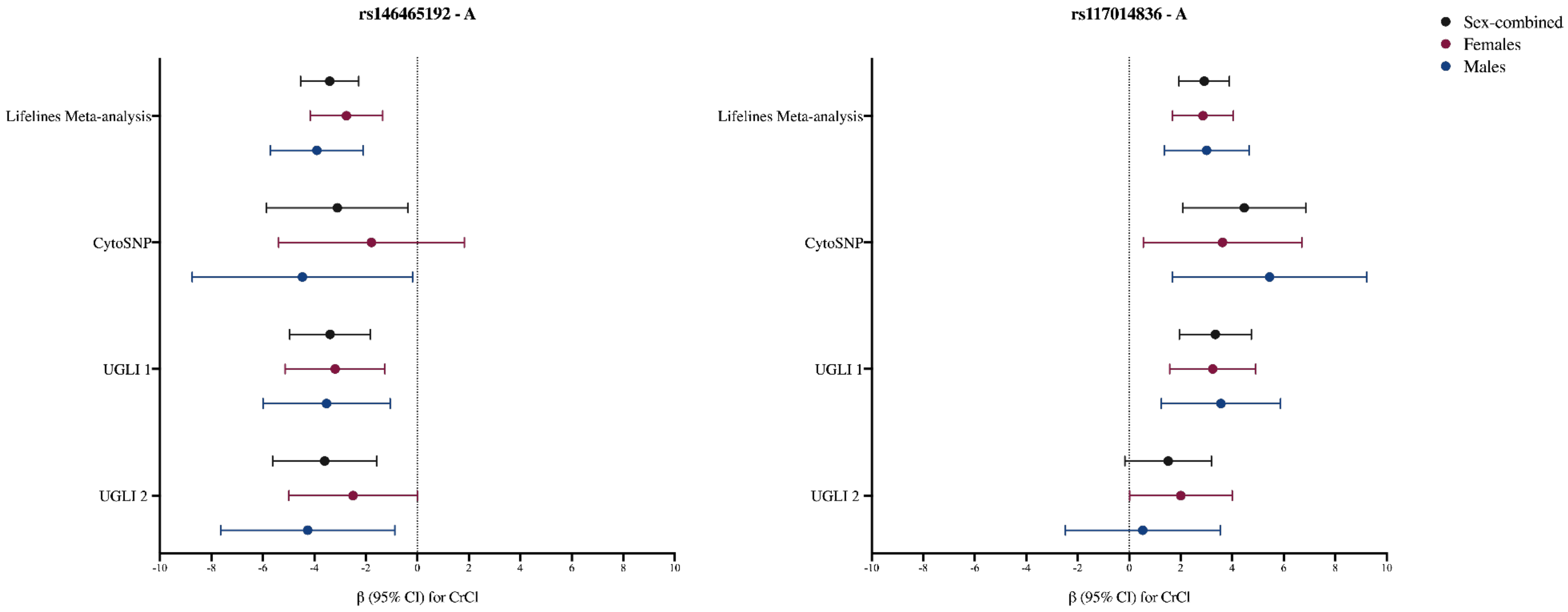

Supplementary Figure S7. Forest plot of CrCl association estimates for the two novel SNPs across the GWAS meta-analysis, CytoSNP, UGLI1, and UGLI2. rs146465192: $I^2$ = 0% for total, female, and male; rs117014836: $I^2$ = 57.3% for total, 0% for female, and 55.15% for male.

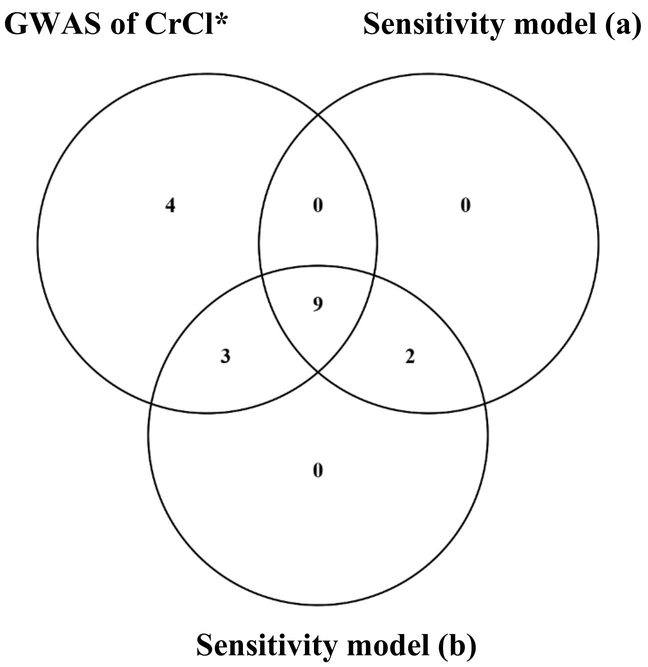

Supplementary Figure S8. Venn diagram of overlapping loci across GWAS models. The Figure shows the number of loci shared between different GWAS models in the sex-combined analyses. *Primary model: CrCl corrected for Body Surface Area (BSA); model: CrCl = SNPs + age + sex + 20 PC. Sensitivity model (a): CrCl unadjusted for BSA; model: CrCl = SNPs + age + sex + 20 PCs. Sensitivity model (b): CrCl unadjusted for BSA, including height as a covariate; model: CrCl = SNPs + age + sex + height + 20 PCs.

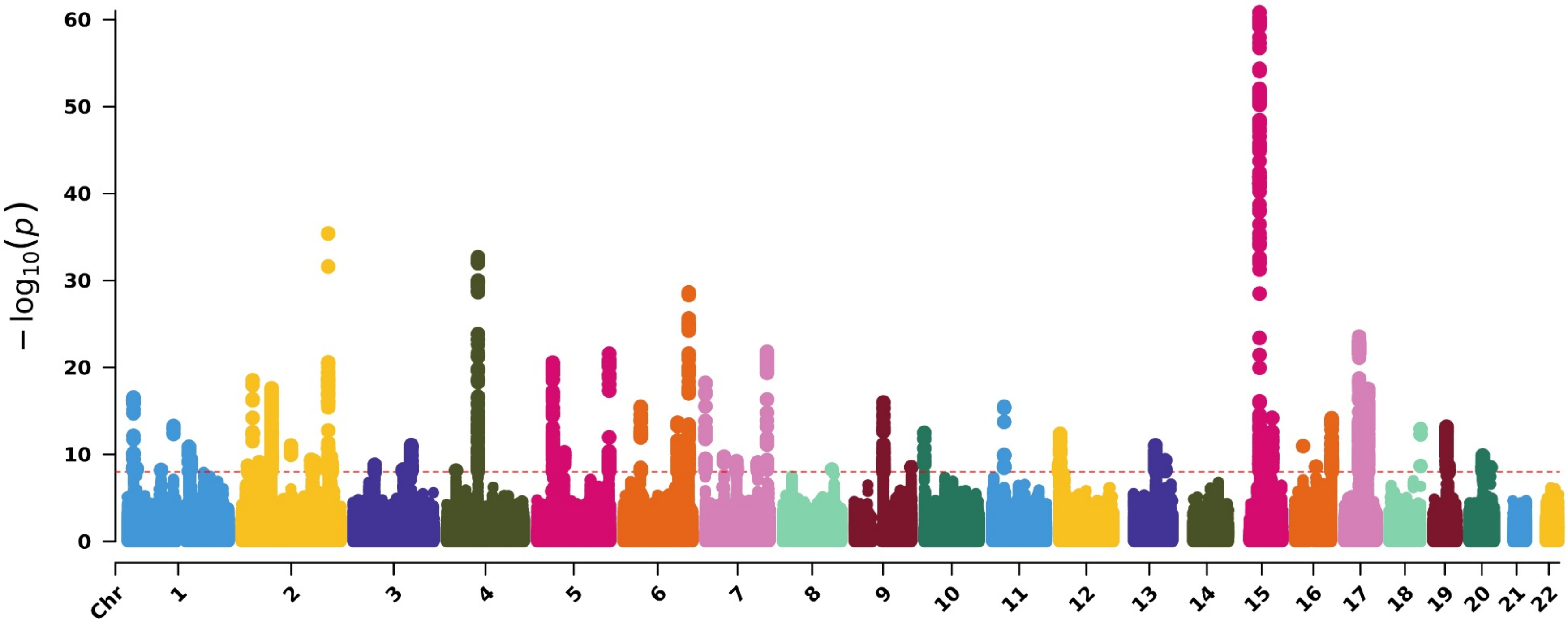

Supplementary Figure S9. Manhattan plot for eGFRcrea GWAS results. Manhattan plot shows –log10 association P value for the genetic effect on eGFRcrea by chromosomal base position (GRCh37). The red line indicates the threshold for genome-wide significance ($p < 5 \times 10^{-8}$). Models adjusted for age, sex and 20 principal components (PCs)

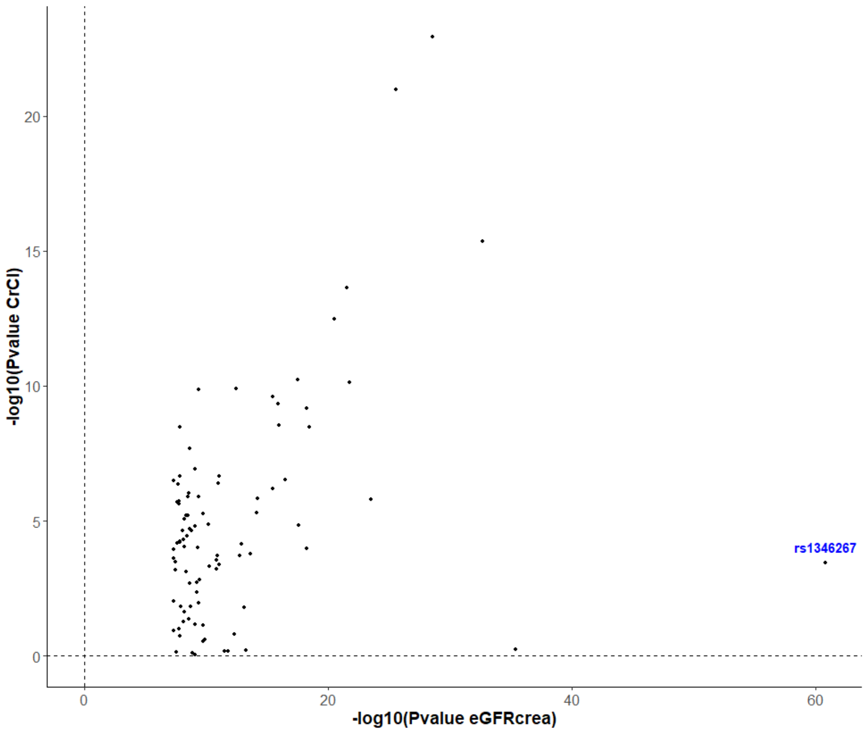

Supplementary Figure S10. Scatter plot of P-values for lead eGFRcrea SNPs versus CrCl GWAS. The plot displays $-\log_{10}$(P) values for lead SNPs identified in the eGFRcrea GWAS compared to their corresponding $-\log_{10}$(P) values in the CrCl GWAS. The highlighted SNP corresponds to the top eGFRcrea-associated variant ($P = 1.53 \times 10^{-61}$).

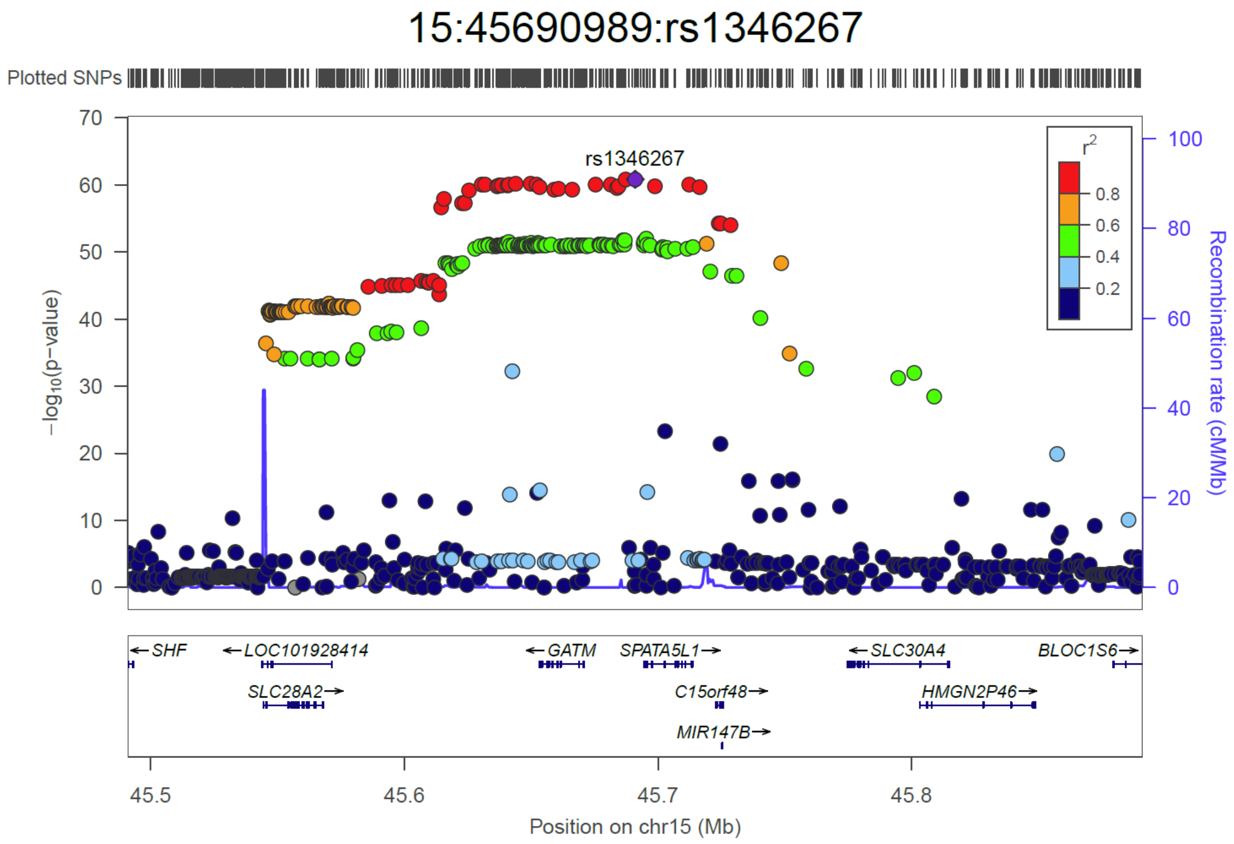

Supplementary Figure S11. Regional plot for the top eGFRcrea hit rs1346267. $r^2$ values are from hg19 / 1000 Genomes EUR reference samples. Plots were generated with LocusZoom (http://csg.sph.umich.edu/locuszoom/).

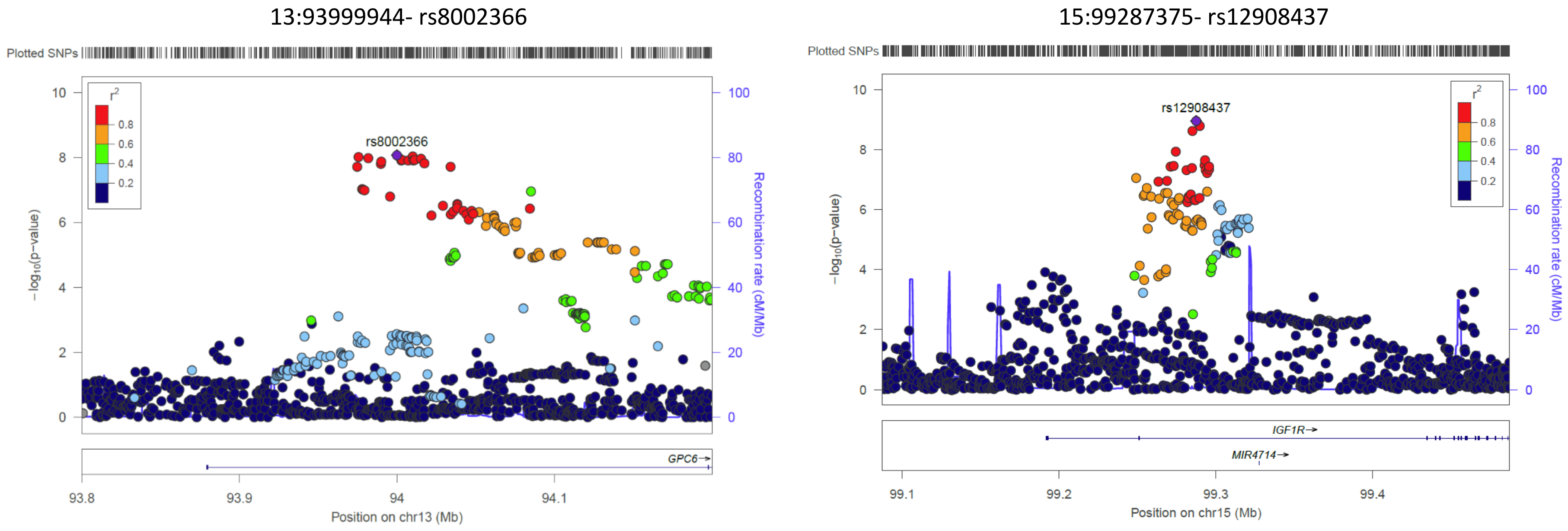

Supplementary Figure S12. Regional plots for unique female-specific SNPs rs8002366 and rs12908437. $r^2$ values are from hg19 / 1000 Genomes EUR reference samples. Plots were generated with LocusZoom (http://csg.sph.umich.edu/locuszoom/).

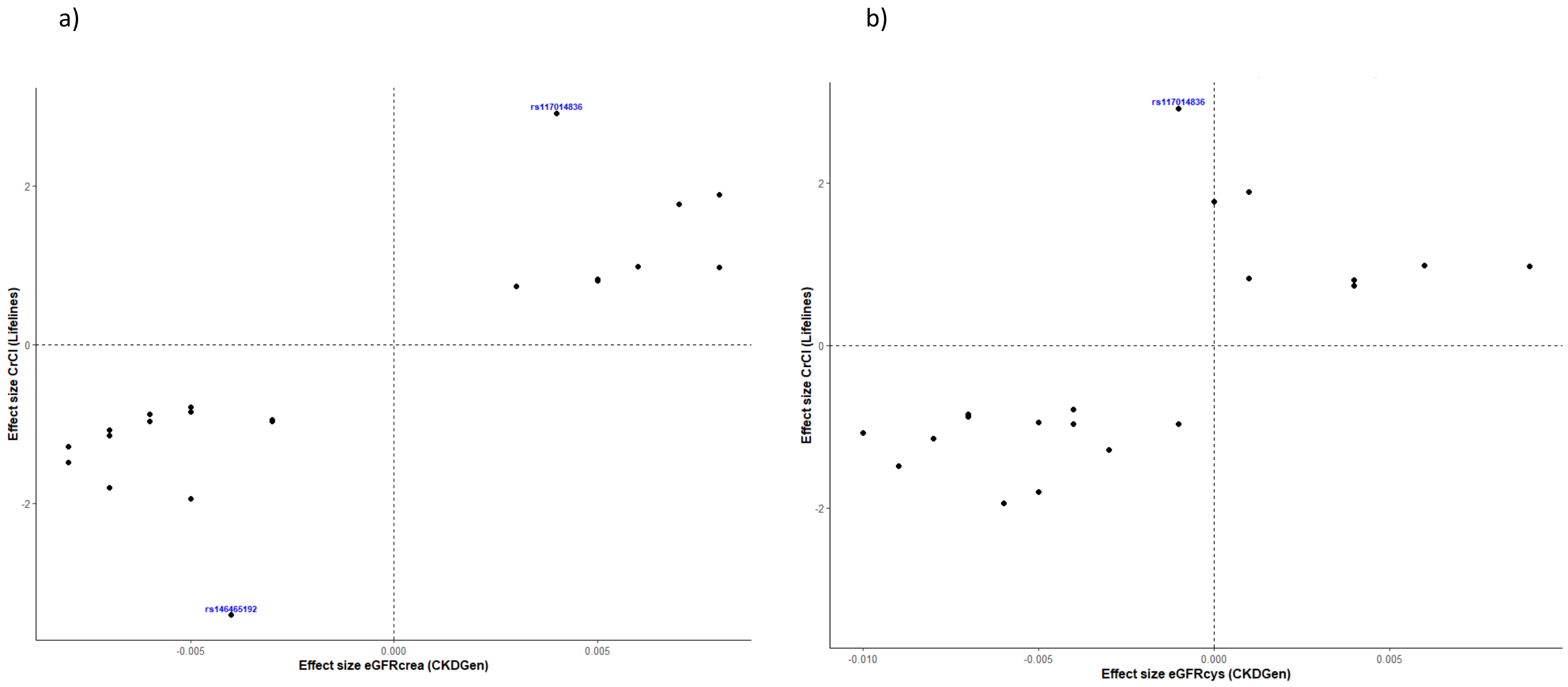

Supplementary Figure S13. Scatter plot of effect sizes for lead CrCl SNPs in Lifelines compared with eGFRcrea (a) and eGFRcys (b) in CKDGen. The two novel SNPs are highlighted in blue. Numerical values are provided in Table 1, Supplementary Data S16, and Supplementary Data S18. The SNP rs146465192 was not present in the eGFRcys summary statistics.

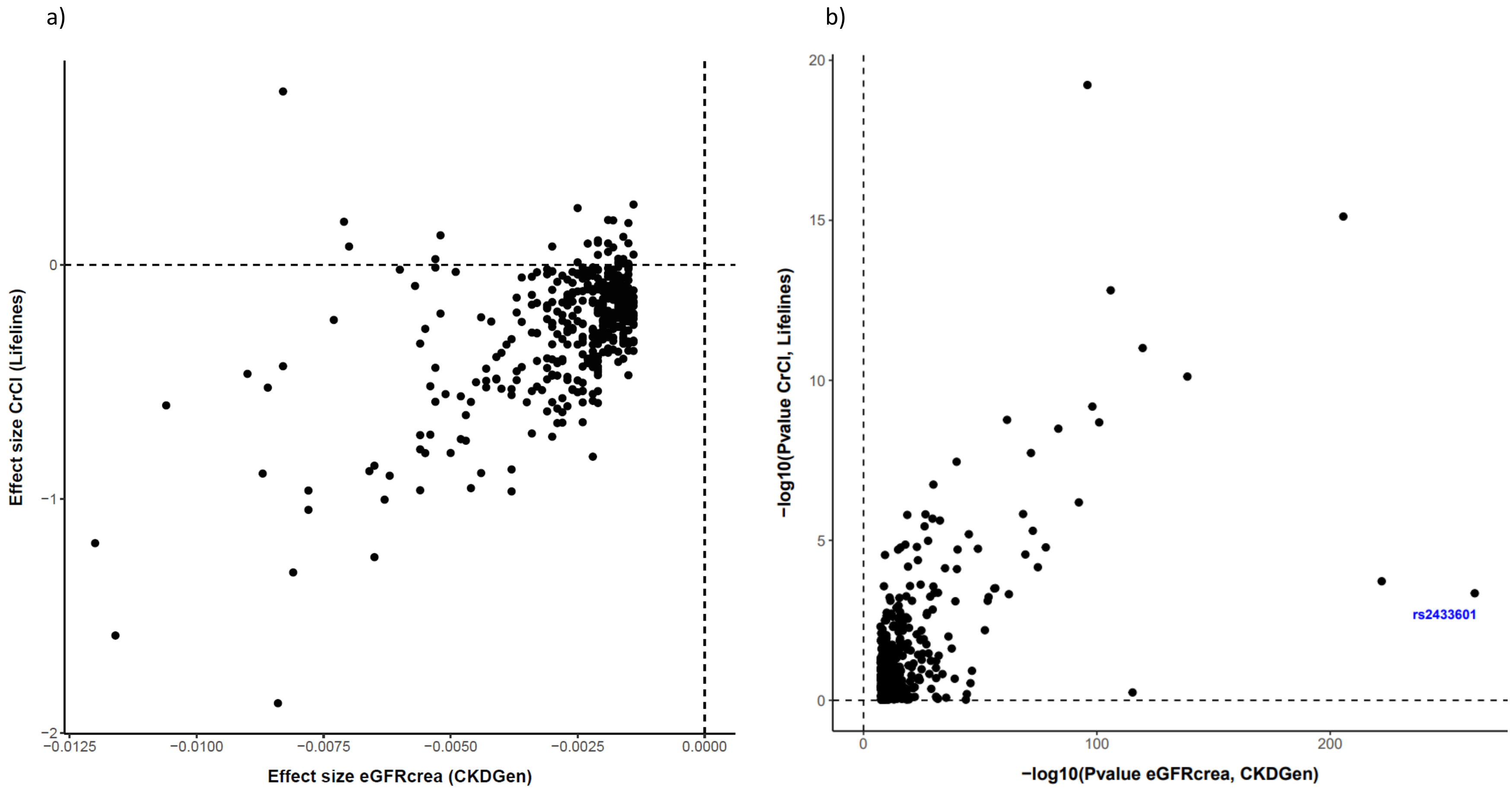

Supplementary Figure S14. Scatter plot of effect sizes (a) and P values (b) of lead eGFRcrea SNPs in CKDGen compared to CrCl in Lifelines. Numerical values are provided in Supplementary Data S19. The strongest eGFRcrea SNP in CKDGen (rs2433601 in the *SPATA5L1* locus) is highlighted in blue $P = 1.13 \times 10^{-262}$.

a)

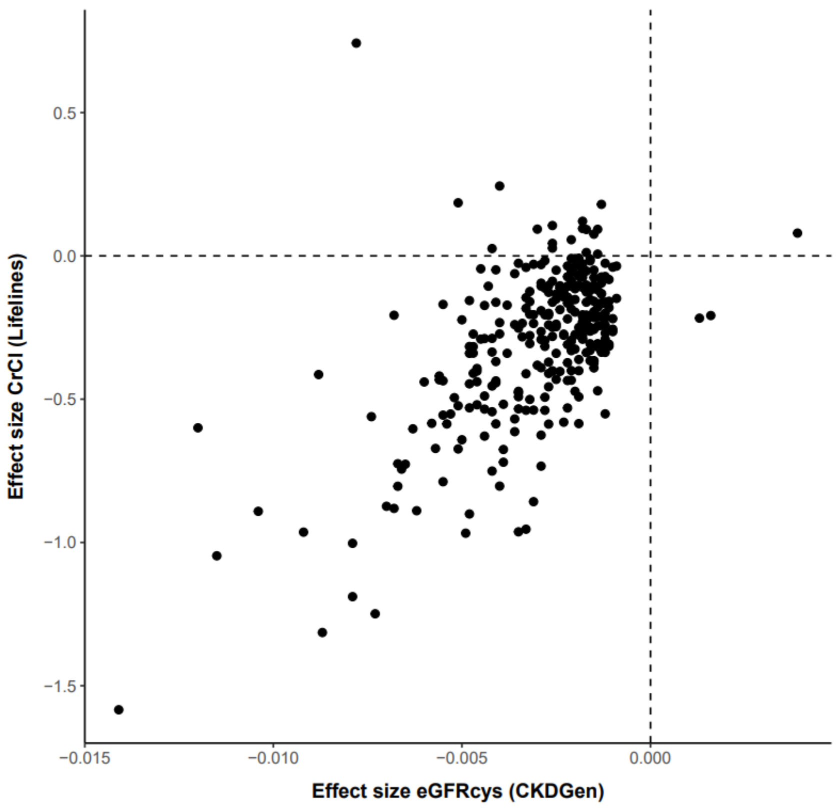

b)

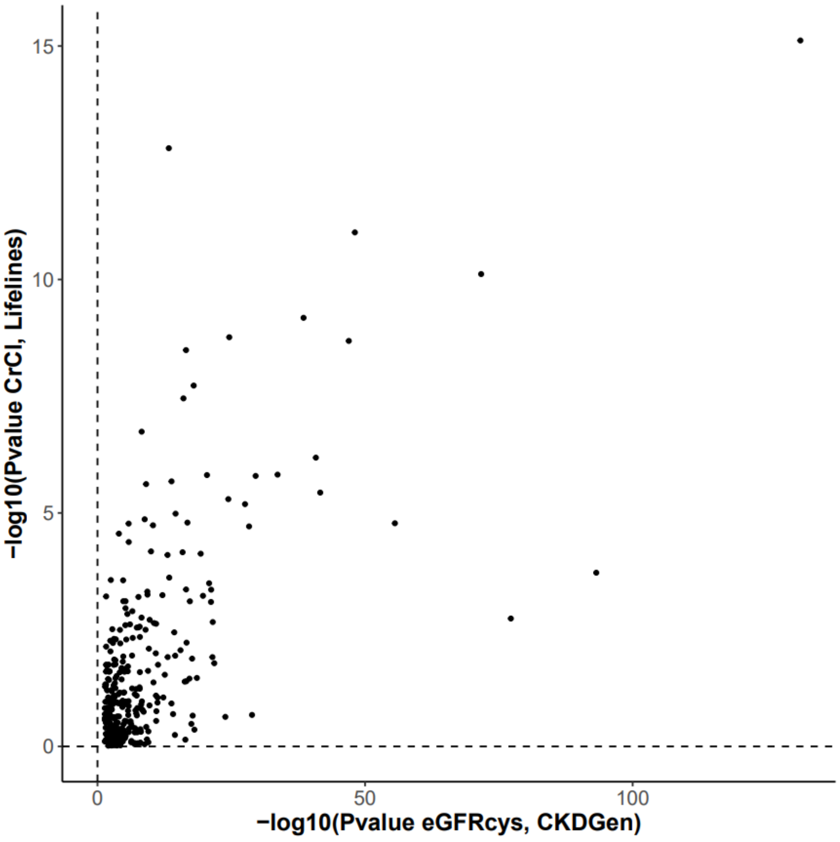

Supplementary Figure S15. Scatter plot of effect sizes (a) and P values (b) of lead eGFRcys SNPs in CKDGen compared to CrCl in Lifelines. Numerical values are provided in Supplementary Data S19.

a.

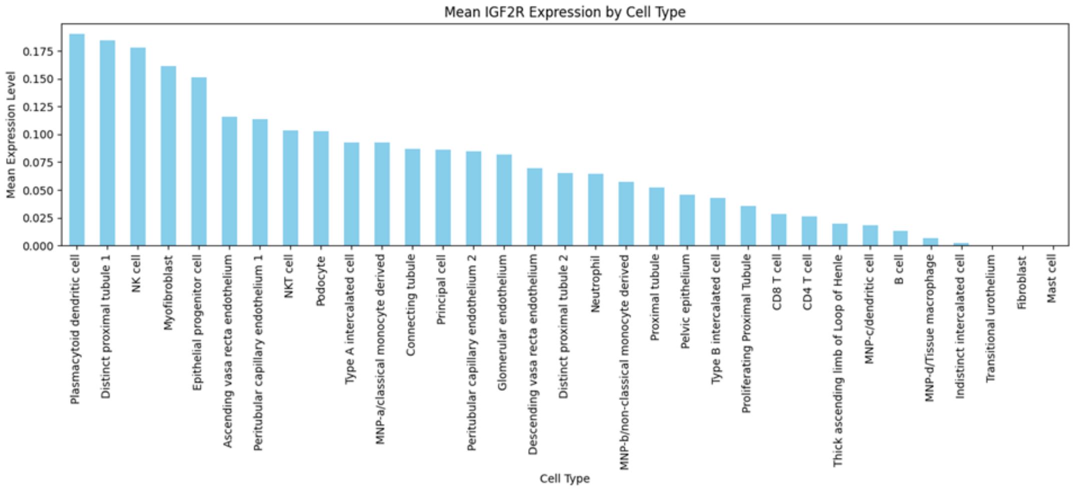

b.

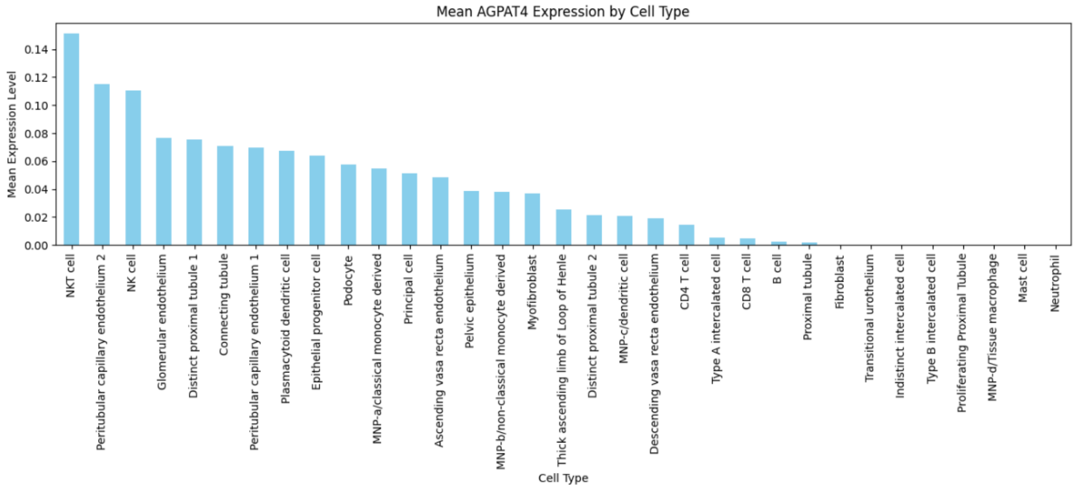

Supplementary Figure S16. Mean IGF2R and AGPAT4 expression across cell types. This figure shows the mean *IGF2R* (a) and *AGPAT4* (b) expression levels across different cell types, including outlier cells.

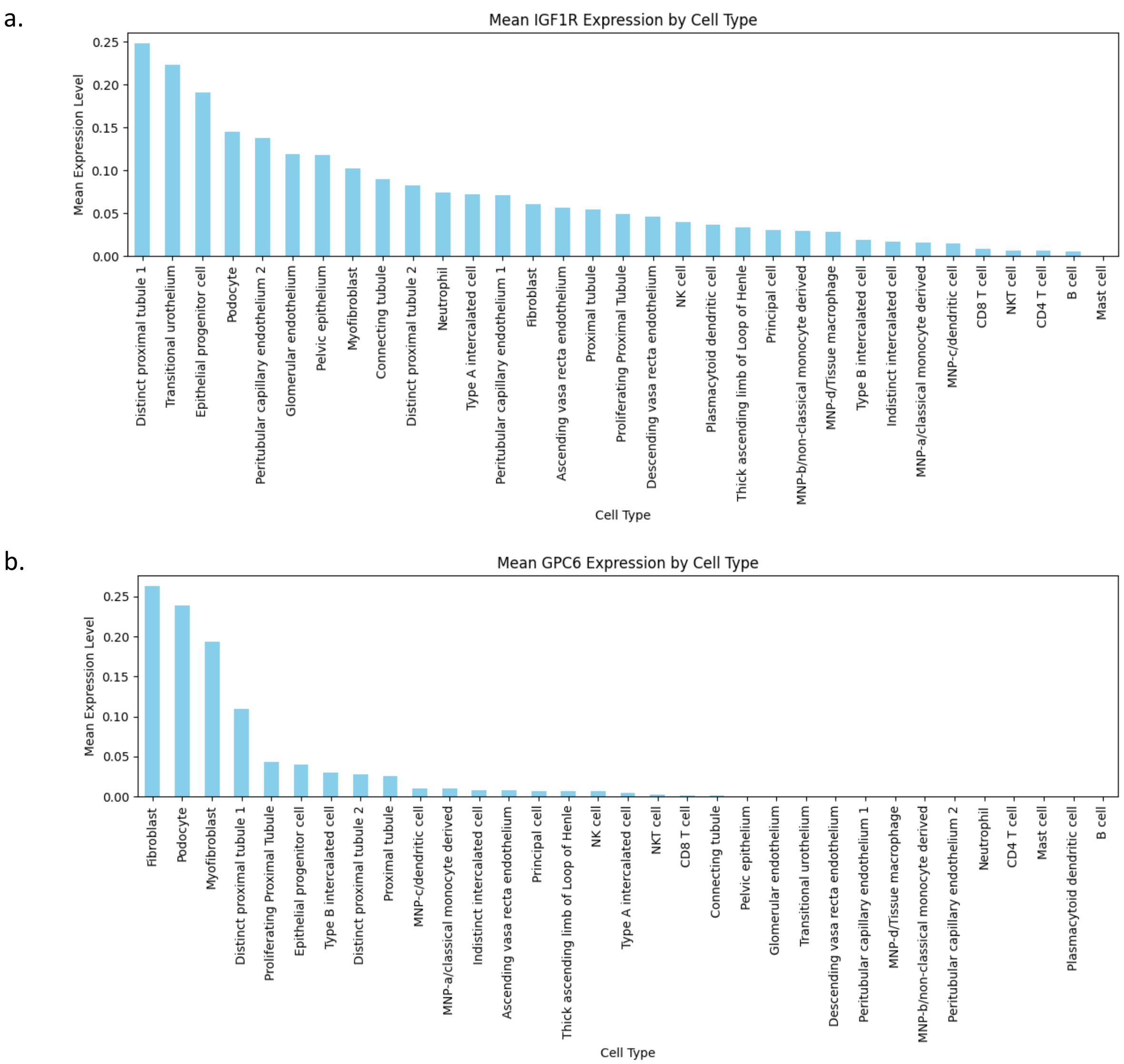

Supplementary Figure S17. Mean IGF1R and GPC6 expression across cell types. This figure shows the mean *IGF1R* (a) and *GPC6* (b) expression levels across different cell types, including outlier cells.

a) b)

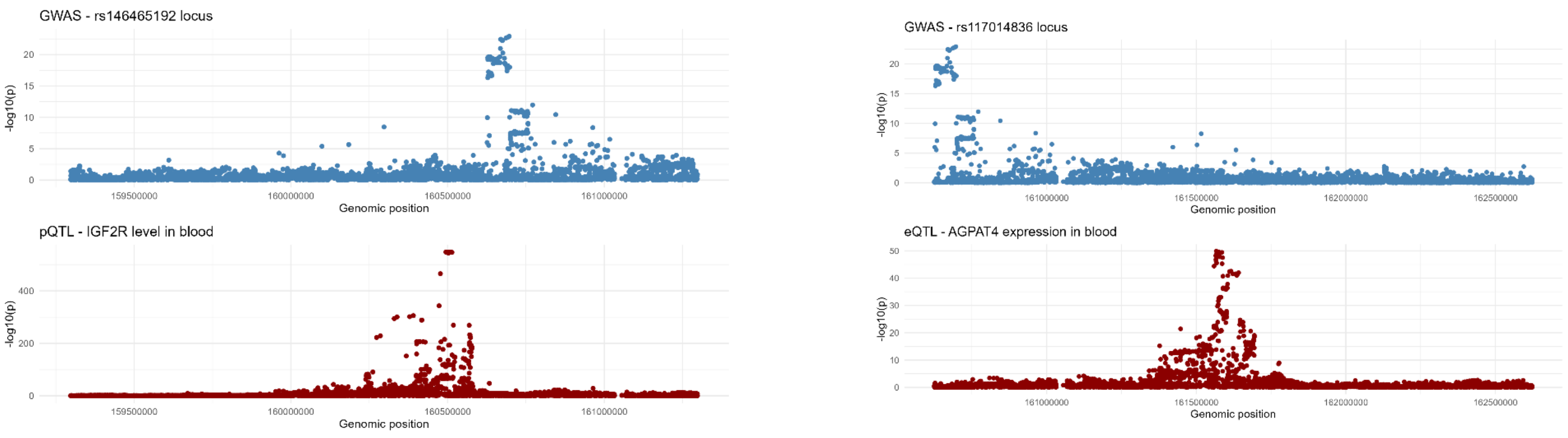

Supplementary Figure S18. Colocalization results for the two novel CrCl-associated SNPs

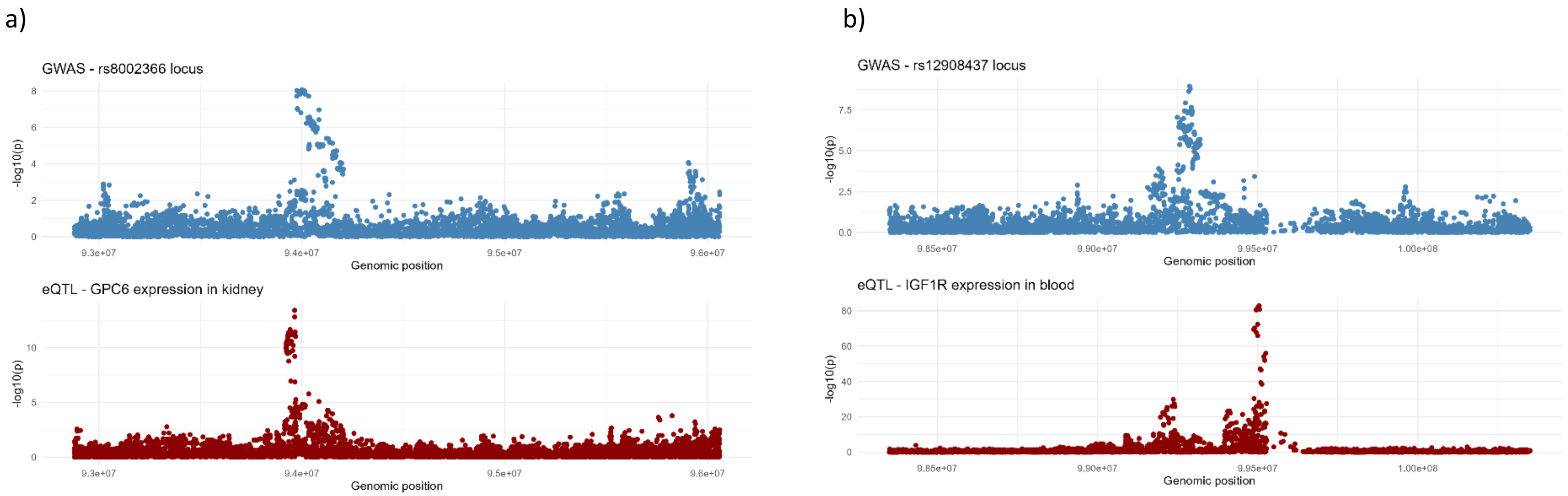

Supplementary Figure S19. Colocalization results for the two female-specific CrCl-associated SNPs

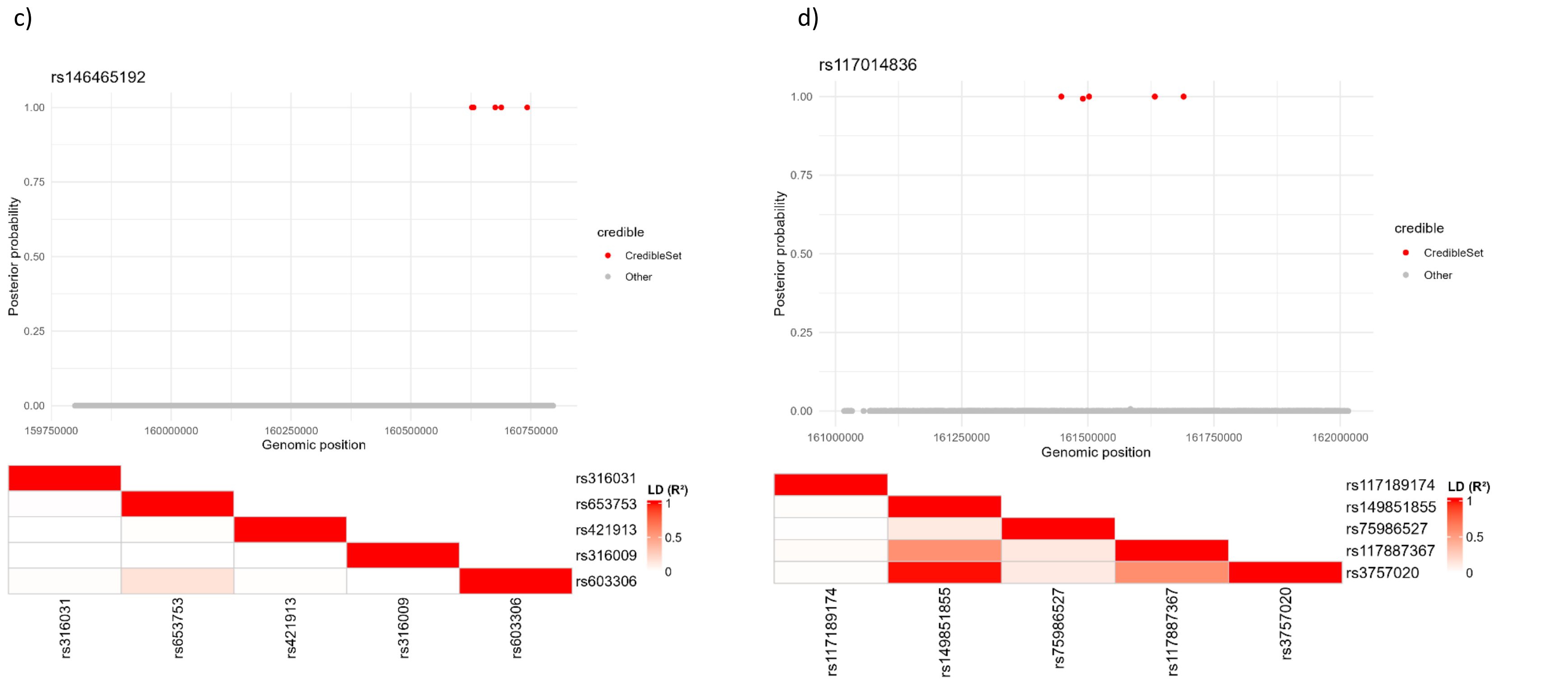

Supplementary Figure S20. Fine-mapping results for the two novel CrCl-associated SNPs

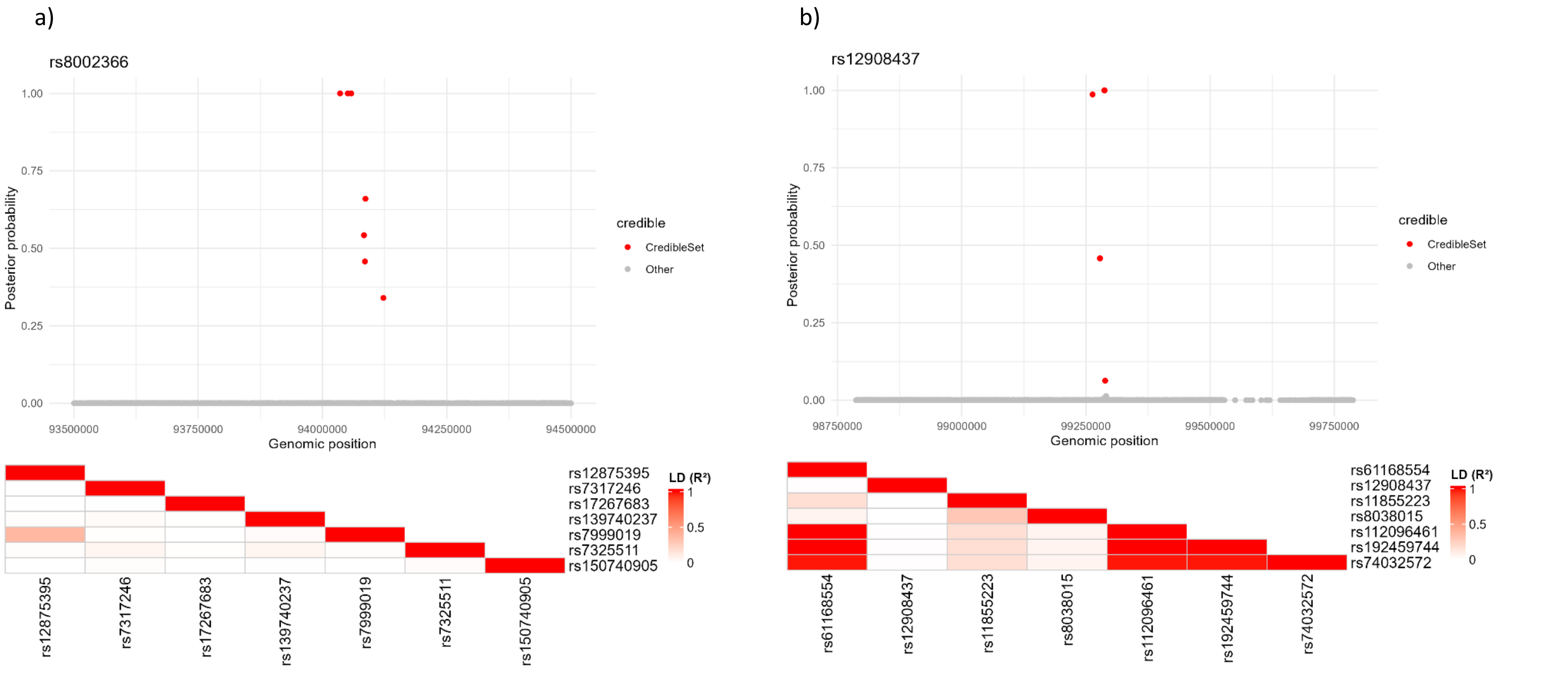

Supplementary Figure S21. Fine-mapping results for the two female-specific CrCl-associated SNPs

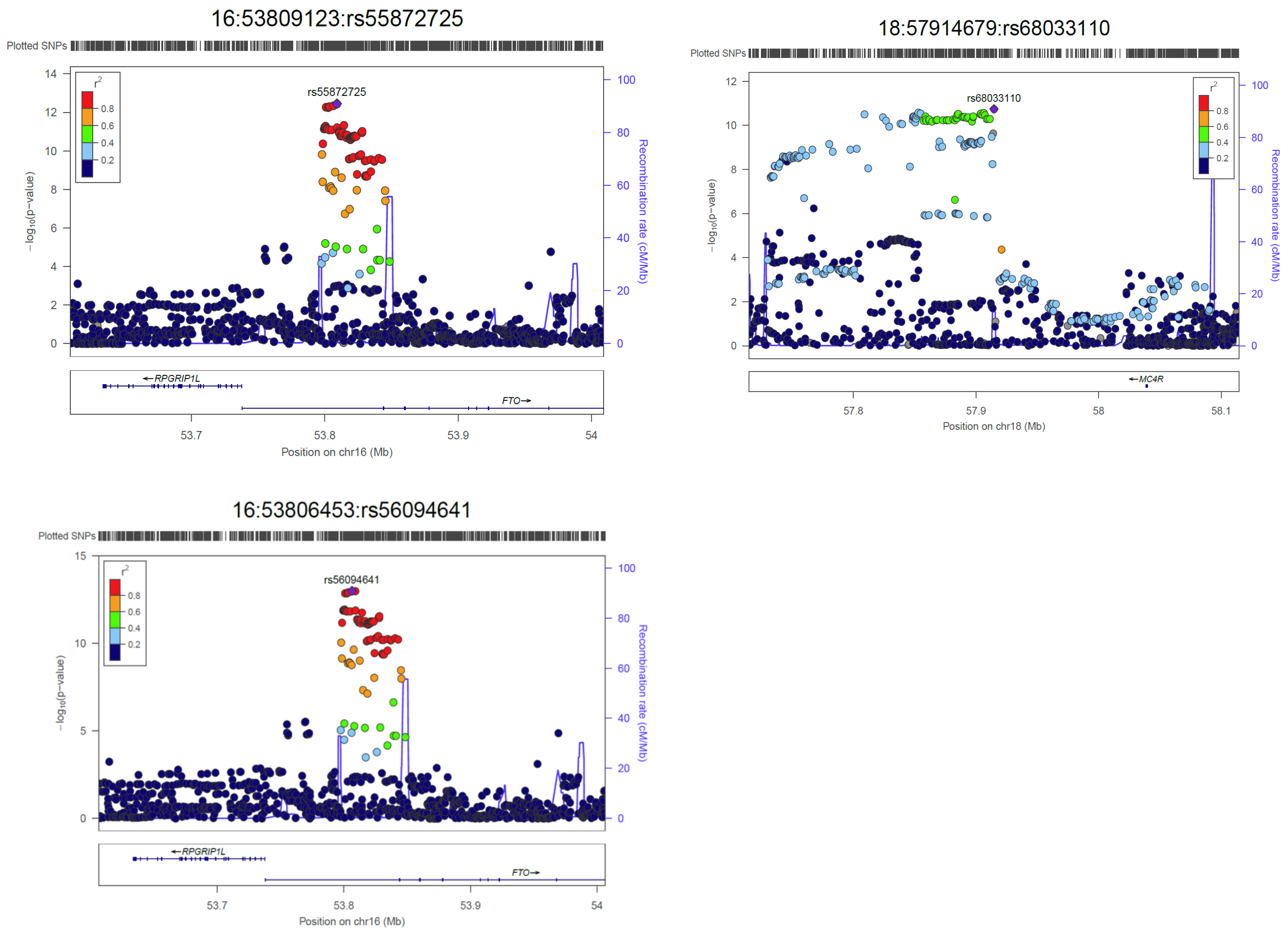

Supplementary Figure S22. Regional plots for three lead SNPs exclusive to CrCl sensitivity GWAS models. $r^2$ values are from hg19 / 1000 Genomes EUR reference samples. Plots were generated with LocusZoom (http://csg.sph.umich.edu/locuszoom/).
